# Modeling the levels, trends, and connectivity of malaria transmission using genomic data from a health facility in Thiès, Senegal

**DOI:** 10.1101/2021.09.17.21263639

**Authors:** Albert Lee, Yaye Die Ndiaye, Aida Badiane, Awa Deme, Rachel F. Daniels, Stephen F. Schaffner, Fatou Ba Fall, Médoune Ndiop, Alioune Badara Gueye, Ibrahima Diallo, Katherine E. Battle, Edward A. Wenger, Caitlin A. Bever, Doudou Sene, Bronwyn MacInnis, Dyann F. Wirth, Daouda Ndiaye, Daniel L. Hartl, Sarah K. Volkman, Joshua L. Proctor

## Abstract

Molecular data and analysis outputs are being integrated into malaria surveillance efforts to provide valuable programmatic insights for national malaria control programs (NMCPs). A plethora of studies from diverse geographies have demonstrated that malaria parasite genetic data can be an important tool for drug resistance monitoring, species identification, outbreak analysis, and transmission characterization. Despite many successful research efforts, there are still important knowledge gaps hindering practical translation of each of these use cases for NMCPs. Here, we leverage epidemiological modeling and time-series data of 2035 genetic sequences collected in Thiès, Senegal from 2006-2018 to provide a quantitative and setting-specific assessment of the levels, trends, and connectivity of malaria transmission. We also identify the genetic features that are the most informative for inferring transmission in Thiès, such as the fraction of the population with multiple infections and the persistence of parasite lineages across multiple transmission seasons. The model fitting and uncertainty quantification framework also reveals a significant decrease in the level of malaria transmission around 2013. This difference coincides with a large-scale drought and bed net campaign by the NMCP and USAID and is independently corroborated by geo-spatial models of incidence in Thiès. We find that genetically identical samples are more likely to be geographically clustered even at the neighborhood scale; and moreover, these lineages propagate non-randomly around the city. Our approach and results provide quantitative guidance for the interpretation of malaria parasite genetic data from Thiès, Senegal and indicates the value of increased malaria genomic surveillance for NMCPs.

## Introduction

Malaria parasite genetic data is poised to be an important tool for national malaria control programs (NMCPs) to better characterize transmission, optimize surveillance programs, design intervention efforts, and ultimately decrease global disease burden. Over the past two decades, research investigations have provided substantial evidence around the programmatic value of genetic data for monitoring the spread of parasites that have genetic markers associated with drug resistance [1, 2, 3] and diagnostic failure [4]. Integration of molecular sampling and data collection into NMCP strategies has already started [5, 6] and is concurrently being considered for incorporation into formal guidance for routine malaria surveillance by the World Health Organization (WHO) [7, 8]. Despite the clear value that molecular data has for these use-cases, there are still substantial gaps in understanding and characterizing the link between parasite genetic data and malaria transmission, and therefore the implications for malaria control strategies. Here, we leverage mathematical modeling and genetic data collected in Thiès, Senegal from 2006-2018 to infer the levels, trends, and connectivity of malaria, quantify the usefulness of genetic features and their links to transmission, and highlight the need for additional studies.

Malaria parasite molecular data have been utilized for a wide set of research and programmatic questions beyond drug and diagnostic resistance detection. Molecular data enables the identification of different malaria species [9, 10] providing especially helpful information in regions where malaria infections were either absent or considered completely imported [11]. An extensive library of parasite genetic samples can also help researchers and programmatic investigators during an outbreak to help determine the geographic origin, rate of importation, and subsequent level of local transmission [12, 13, 11]. More broadly, genetic similarity between samples has been used to identify the parasite connectivity between human populations [14, 15, 16, 17, 18, 19], which may provide a pragmatic and cost-effective tool to inform control programs as they plan local elimination efforts [20]. In addition, malaria parasite genetic data has also been linked to helping assess transmission levels and trends [21] using approaches from population genetics [20] and epidemiological modeling [22, 23, 24, 25, 26].

Mathematical modeling has been a valuable tool to investigate the links between transmission characteristics and features extracted from the malaria parasite genetic data. Early efforts by Daniels et al. [22] demonstrated that a parsimonious, malaria genomic model could be calibrated to genetic data and detect changes in transmission; that work provided further evidence toward the hypothesis that genetic features such as the proportion of the population with multiple infections and the persistence of clones in a population can be linked to transmission [27, 28]. Subsequent modeling efforts have added details of parasite life cycles and genomic evolution, including processes such as meiotic recombination and superinfection [29, 23, 24, 26, 25]. These efforts have explored the qualitative relationships between genetic features, their relationship to transmission, the impact of sampling, and compared their findings to malaria genomic datasets [23, 24]. Here, we present an agent based generalization of the malaria epidemiological genomic model from [22] allowing for better strain tracking, more realistic meiotic recombination processes, and genomic reservoir initialization. We directly fit the model to genetic samples collected from 2006 to 2018 allowing for a quantitative assessment of the levels and trends of transmission over time and specific investigation of the most informative genetic features for Thiès.

We also leverage probabilistic movement models to investigate parasite connectivity within Thiès. Features derived from genetic data, such as identity-by-descent for whole genome sequencing and identity-by-state for single-nucleotide polymorphism (SNP) panels, along with associated metadata have helped identify parasite connectivity across tens and hundreds of kilometers [14, 15, 16]. Innovative efforts have also integrated complementary data from mobile-phone data and travel surveys with malaria parasite genetics [17]. In this work, we demonstrate that malaria parasite genetic data can identify connectivity at a small geographic scale; we utilize spatial models of human movement that have been shown, for various pathogens, to be effective for characterizing genetic linkages [30, 31]. The modeling investigation in this article leads to a better understanding of the levels, trends, and connectivity of malaria in Thiès, Senegal, and can inform decision-making for interventions in local settings.

## 1 Results

We analyzed 2035 samples comprising 13 years of genomic data from *Plasmodium falciparum* infections in Thiès, Senegal. We genotyped 686 samples for 24 unlinked SNPs (a molecular barcode) from 2014 to 2018 in combination with 1349 previously published barcodes [22, 27]. Figure 1.A illustrates the number of samples collected each year and the proportion of samples with multiple genomes (polygenomic infections). Among samples with a single genome (monogenomic infections), 173 distinct barcodes appeared more than once. Figure 1.B shows the persistence of monogenomic infections by displaying the most frequently observed lineages. Some barcodes from 2014-2018 were continuations of lineages from 2006-2013 (26 samples); also, we observed lineages in the new dataset that were not previously observed from 2006-2013 (69 samples).

**Figure 1:**
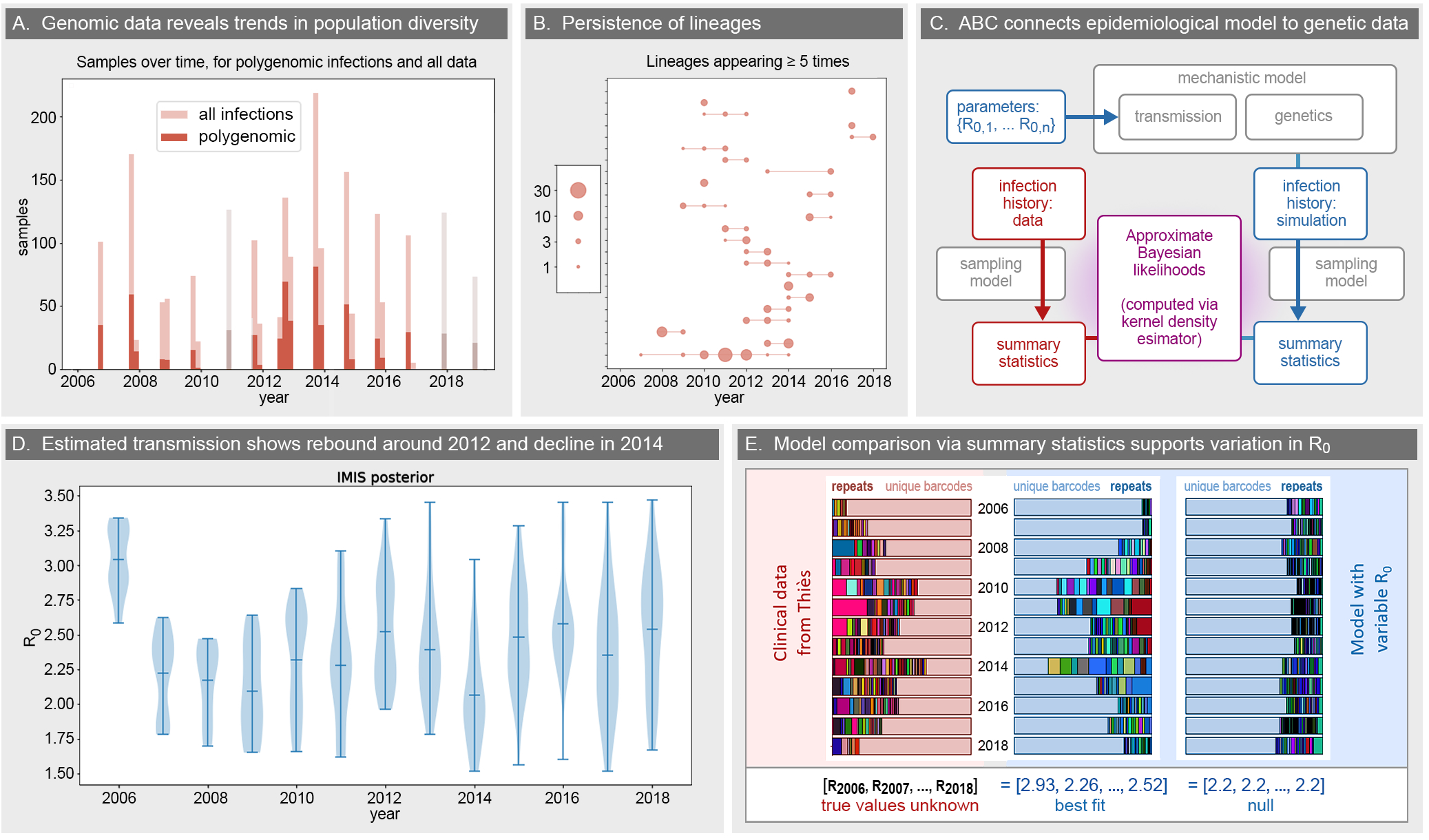
Overview of the malaria genomic model. **A**. Number of clinical infections (samples) used in this study, counted in two-month bins. The number of polygenomic samples are darker shaded. Samples for which only the year is known are shown in gray. **B**. Many barcodes form lineages, and some persist for multiple years (lineages with at least 5 observations are shown above). The longest lineage is observed from 2007 to 2014. **C**. Conceptual schematic of the dynamical malaria transmission model and passive sampling model (gray), generating summary statistics from the model (blue) and data (red), and the comparison framework using approximate Bayesian computation (ABC) and kernel density estimators (magenta). **D**. The posterior distribution from incremental mixture importance sampling calibration results for the input parameter *R*_0_, specifying the reproductive number for each year. The mean values and 95% credible intervals are shown as horizontal ticks, and the uncertainty is visualized by the length of the violin plot. **E**. An illustration to represent summary statistics for ABC, computed for both the data (red, left), a stochastic replicate of the best-fit model (blue, middle), and a stochastic replicate of a null model with no variation in *R*_0_ (blue, right).

### 1.1 Inferred levels of malaria transmission highlight a decrease in transmission from 2012 to 2014

Transmission levels were inferred from an agent-based genetic model using approximate Bayesian computation (ABC) to calibrate *R*_0_ (Fig. 1.C). The estimated transmission decreased substantially from 2006 to 2008 and eventually rebounded to higher levels in 2012. Figure 1.D shows the mean and posterior distribution of the reproductive number fit; see Supplement Table S1 for a more detailed description of the kernel density estimates of *R*_0_. The joint posterior distribution over *R*_0_ parameters indicates that the levels of transmission have likely changed from 2006 to 2018 (we estimate that *P* (Δ*R*_0_ < 2.5) < 10^−5^ where Δ*R*_0_ is the total absolute change in *R*_0_ over a simulation). A similar result was found by Daniels et al. [22] from 2006 to 2013 (Supplement Figure S4). Figure 1.E provides a visual comparison of a portion of the summary statistics derived from genetic data between the clinical data (left, red), a single simulation generated with the best fit model, and a single simulation generated from a flat transmission level of 2.2. The relative decrease in transmission between 2006 and 2010 is approximately 30%. The decrease occurred over a two year period followed by a three year stable level of transmission with a gradual rebound to higher levels of transmission around 2012.

The estimated transmission level sharply decreased from 2012 to 2014. From the marginal joint posterior, the probability that *R*_0_ decreased from 2012 to 2014 is 0.92, and the probability that it decreased by at least 10% is 0.70. The inferred change in transmission is also directly apparent by evaluating the summary statistics of genetic data; similar trends can be seen visually in Figure 1.E. Note that the proportion of unique barcodes versus repeats also undergoes a rebound from 2010 to 2012 and drops in 2014. The inferred transmission levels and trends are consistent for different calibration methods; we find qualitative agreement when varying the underlying assumptions of the mechanistic model, the sampling of infections in the model, and the likelihood for the ABC model fitting procedure; see Supplement §1 for more details.

### 1.2 Proportion of polygenomic barcodes, unique barcodes, and number of repeated strains are the most informative summary statistics for inferring modeled transmission levels

Among 58 summary statistics we analyzed (Supplement § 3.A), the yearly population fraction of samples that are polygenomic (*f*_*P,y*_) are the most informative summary statistics for fitting the malaria genomic model to barcode data from Thiès, Senegal (Supplement § 3.C). Other statistics that are informative include the yearly proportion of barcodes that are unique (*f*_*U,y*_) and the number of barcode strains that are repeated twice (*n*_2,*y*_). This result is consistent whether evaluating the correlation between model parameter estimates and associated summary statistics or a feature-selection regression (Methods and Supplement § 3.C). For example, the Spearman correlation between *f*_*P,y*_ and *R*_0,*y*_ within the same year can be up to 0.93. This is consistent with dimensional reduction via the least absolute shrinkage and selection operator (LASSO), which converges upon *f*_*P,y*_ as the most predictive statistic of any given year *y* (Supplement §3.C and Tables S2, S3). Furthermore, both approaches indicate that these informative genetic summary statistics are also correlated with the previous and following year’s transmission levels, suggesting their potential for detecting past trends or inferring those of subsequent seasons.

Additionally, the collection days associated with the sampled infections provide an opportunity to construct within-season summary statistics based on polygenomic samples. However, we do not find a consistent empirical trend for the within-season polygenomic fraction (Supplement Figure S9). Owing to comparatively low sample numbers and the variability of start and end times of sample collection each season, we observe large uncertainty intervals at the beginning and end of seasonal trends (Supplement §3.A.1). The average seasonal behavior aggregated over all years indicates a declining trend for the Thiès data, whereas simulations indicate the opposite trend (Supplement Figure S10). Nevertheless, the polygenomic fraction is an informative summary statistic on yearly timescales, and future studies with more systematic sampling across seasons may reveal more granular temporal genetic indicators of transmission properties.

### 1.3 Identical barcodes are locally clustered within Thiès

The distribution of pairwise distances between identical barcodes within a single year (3000 pairs) shows a majority occurring within 3 km of each other (Figure 2.A). The clonal pairwise distance distribution is statistically different from the distribution of distances between all non-clones within a year (*t*-test *p*-value < 3.6 × 10^−6^, with 374714 non-clonal pairs) and a null distribution derived from all 2035^2^ pairs (*t*-test *p*-value < 3.4 *×* 10^−6^). Using only pairs from the same year, a bootstrapping analysis of the distances between randomized sets of clones, non-clones, and randomly permuted sample origins representing a null distribution, also confirms the clonal distribution to be significantly different from non-clones. We find that both the means and medians of the pairwise distances from the bootstrapping analysis are significantly smaller than those for the null model (Figure 2.B, and Supplement Fig. S12). The distribution of distances between non-clones is centered around 2.5 km, the average distance for all samples, and closely resembles the null distribution. A similar finding occurs for distributions using all pairwise distances regardless of the year of collection (Supplement, Fig. S13). We also observe that the distribution of distances between clones suggests more detailed dynamics for individual strains. For example, barcode TACCCCGGTCCAC-CAATAAGACTG appears 60 times over an eight-year period; it is first observed in the Cité Ousmane Ngom neighborhood and qualitatively spreads to more parts of the city (Supplement Fig. S15). Section 1.4 provides a more dynamic interpretation of the appearance and spread of clonal strains.

**Figure 2:**
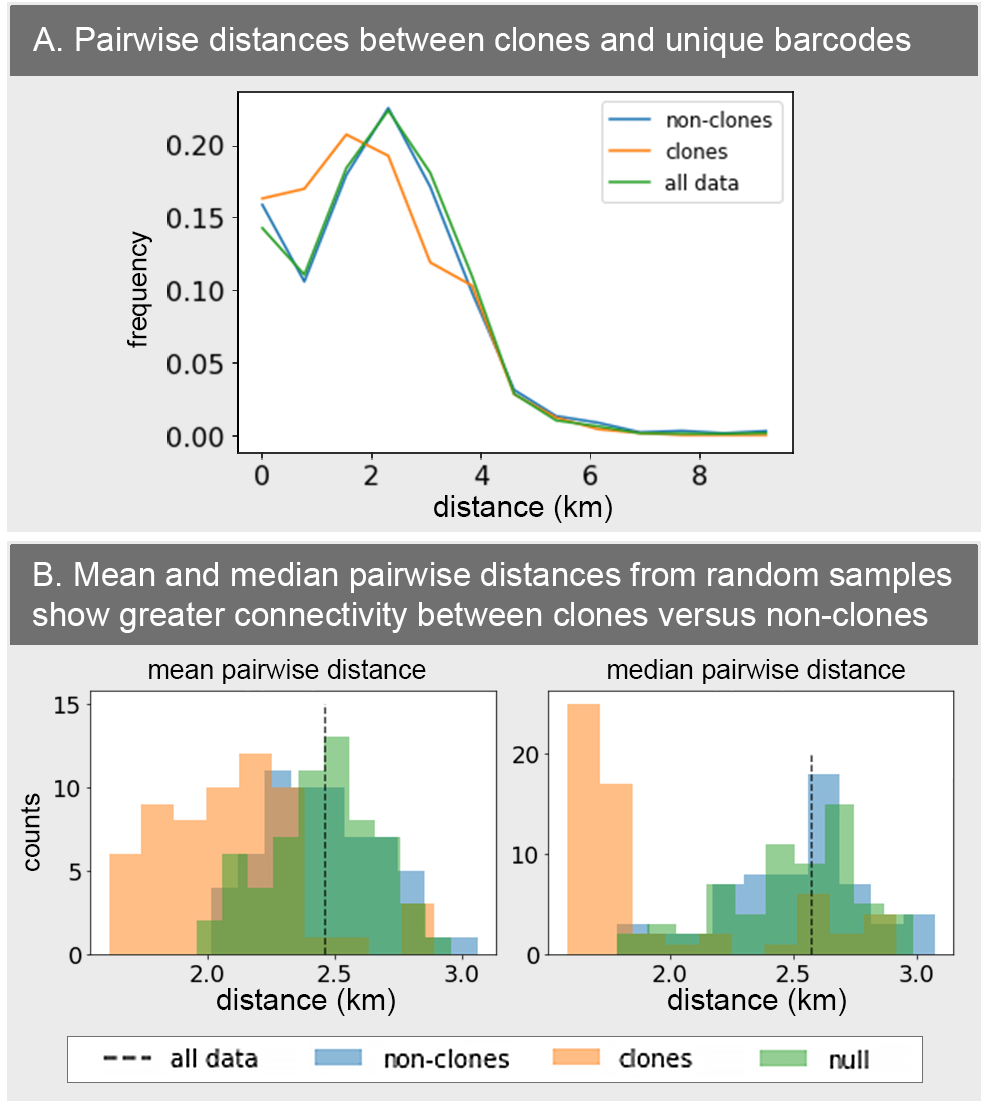
**A**. Distribution of pairwise distances in data, between clones from the same year (orange) and between non-clones from the same year (blue). The distribution for all pairs across all years (green) is provided for comparison. **B**. Distributions of mean and median pairwise distances from randomized subsamples of data, for clones from the same year (orange) and non-clones from the same year (blue). The non-clonal distributions are centered around the mean and median distance for all data (dashed line) and closely resembles a null distribution from permuted samples (green).

### 1.4 A subset of persistent strains propagates non-randomly through the city

The geographic distance between identical barcodes is also correlated with when the samples were collected (Figure 3). For example, Figure 3.A illustrates the appearance of a barcode four times across Thiès during 2013 along with the first principal component corresponding to the direction with the largest variation in space and time (characterized by a slope *s* ≈ 0.008 km/day, Figure 3.B). There is substantial variation across clonal groups, driven by a small number of lineages with some having little to no correlation in space and time (Supplement Figures S16, S17). However, there are 27 persistent strains that exhibit a strong correlation in space and time with an average principal component for these clonal groups corresponding to *s* ≈ 0.021 (km/day) with a standard deviation of *σ*_*s*_ ≈ .003 (km/day). We also compare this observed correlation among persistent barcodes to that of simulations from three spatial dynamic models: two propagation models parameterized with strong local connectivity (one continuous and one discrete) and a null model where barcodes appear randomly in time and space (Fig. 3.C, Supplement §4B). The empirical correlation slope is statistically different from the null model (*p*-value < 10^−3^ for permutation null, *p*-value < 10^−4^ for uniform null); moreover, both propagation models are different from the null (Supplement §4B). These spatiotemporal correlations imply a pattern of local connectivity between clones. Whether this is a signal of a specific transmission mechanism or a consequence of ecological properties remains to be investigated.

**Figure 3:**
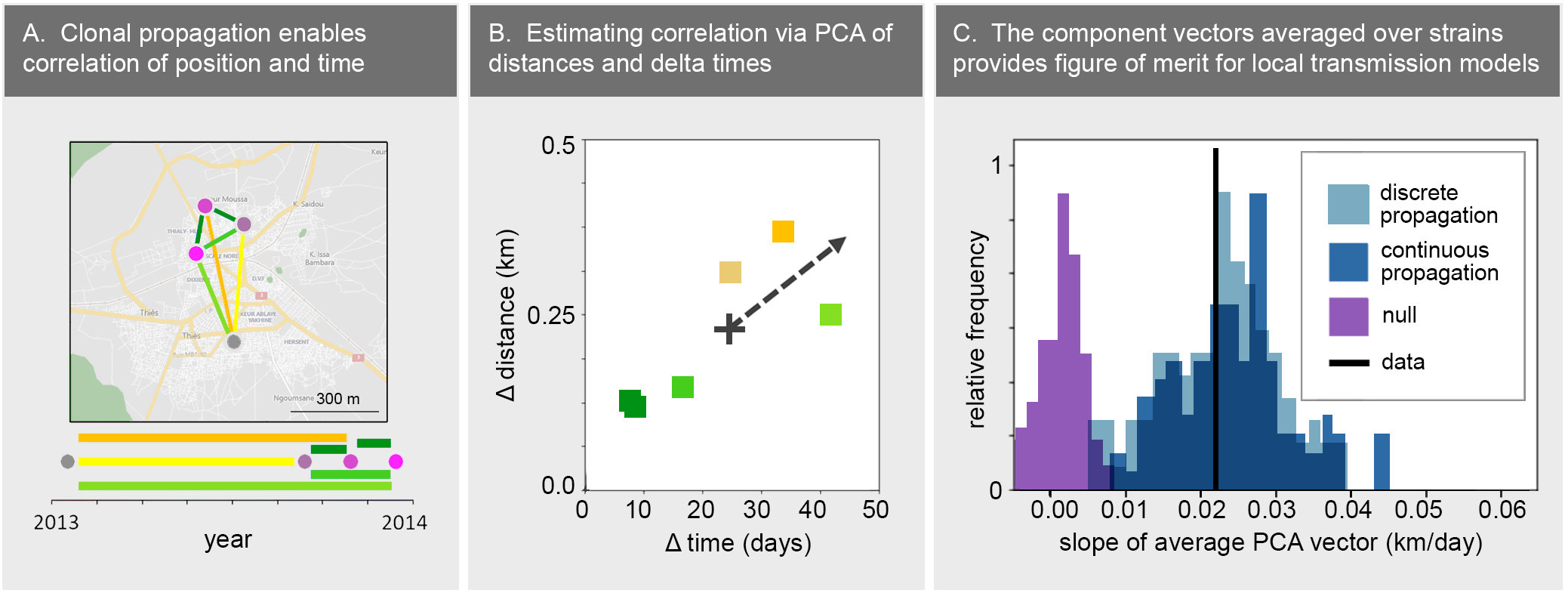
**(A)** For each set of repeated clones, we extract pairwise distances and times between observations. Here, the strain TACCCGGGATCGCACACTAAATTG appears four times at the locations shown on the map and the points on the timeline (in fuchsia). The colored lines indicate the pairwise distances and times elapsed. **(B)** Principal component analysis is used to estimate the correlation between time and distance traveled. The colors of the square data points correspond to the colors of the lines in panel A. The vector 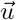 with the larger singular value (black arrow) indicates the direction of greatest variance in parameter space. We let the correlation vector be the average over 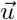 for strains with strong correlations (see Supplement § 4B for details). **(C)** The slope of the correlation vector estimated from Thiès data (black line), compared to the distribution of slopes from a discrete propagation model (light blue), a continuous propagation model (dark blue), and a null model (purple).

## 2 Discussion

We have extended the scope of previous analyses and modeling efforts of malaria transmission and parasite genetics. The malaria genomic epidemiology model is significantly improved from an earlier version [22]; for example, the model now allows for individual-level strain tracking and resolution of multiple infections by individual (Methods and Supplement §2). These innovations enable a more comprehensive investigation into the link between malaria transmission and genetic signals such as complex infections and persistence of specific lineages in a population. We calibrate this model to a complete set of 2035 barcodes across 13 years (§1) to infer transmission levels and trends for Thiès, Senegal from 2006 to 2018. While the inferred transmission levels are consistent with earlier model-based estimates [22] (Supplement Figure S3), our new estimates leverage the full available genetic data across 13 years, such as long-term persistence and emergence of new lineages. Our results provide additional evidence that a relatively small number of genetic samples each year can generate insights into the local levels and trends of malaria transmission, thus augmenting current malaria surveillance efforts.

In more detail, the fitting procedure and concurrent uncertainty quantification also enable insight into the genetic features most informative for inferring the levels and trends of transmission within Thiès. In our region specific calibration, the polygenomic fraction, the proportion of unique barcodes, and number of repeated barcodes are the most informative (§1.2). Three independent methodologies leveraging correlations between genetic features and model parameter fits confirm this result (§1.2 and Supplement §3). This result is consistent with other qualitative malaria parasite genetic modeling efforts [24] and their model output comparison to larger, aggregate datasets, such as pf3k [23]. More broadly, our setting-specific data, model, and inference procedure constitutes a modeling framework to investigate genetic features derived from the data (Fig. 1 and §1.2) and how they are affected by transmission intensity (Supplement §3.C) and variability across stochastic realizations of the model (Supplement §3.C); in addition, these genetic features may be linked to consecutive years suggesting their potential as leading or lagging indicators of transmission (Fig. S2, §S3). Most importantly, though, this Senegal-specific modeling approach can be leveraged to help distill complex genetic data into a key and minimal set of genetic transmission indicators for NMCPs to track and interpret alongside standard clinical indicators.

The new modeling framework may also provide a programmatic interpretation of genetic data relative to transmission and the impact of interventions within Thiès, Senegal. We find a statistically significant decline and rebound of transmission from 2006-2013 similar to Daniels et al. [22], which is correlated with deployment of large-scale malaria interventions over the same period [32]. The inference also identifies a distinctly lower level of transmission in 2013 and 2014 compared to 2012 (Figure 1.E); the posterior distributions, generated by the Bayesian model fitting procedure, indicate the differences are indeed significant (§1.1). The reduced transmission levels in 2013 and 2014 are also observed in recent model-based estimates of malaria prevalence by the Malaria Atlas Project (MAP) for all of Africa [33], which were primarily informed by household survey data and environmental data. In addition, this pattern is confirmed by preliminary efforts by MAP to generate a Senegalese-specific incidence map based on both routine surveillance and household survey data. The timing of this estimated decrease in transmission is correlated with the roll out of free and subsidized long-lasting insecticide-treated bed nets nationwide beginning in 2013 and continuing into 2014 by the Senegal NMCP, supported by USAID through the President’s Malaria Initiative [34]. These associations with the model-based inference support previous assertions that genetic data can reveal changes in transmission [22], however, an indepth understanding of the causal relationship between climate, interventions, transmission, and genetic signals will require an expansion of the current modeling effort and systematically linking clinical, programmatic, environmental, and genetic data by region. Nevertheless, these results suggest that genetic features can reflect decreases in transmission and could provide an evaluation and monitoring tool for programmatic interventions.

Modeling has also shown that malaria genomic data can help reveal the local connectivity of populations at the geographic scale of Thiès, Senegal. We found identical barcodes are more likely to be geographically localized than non-clones (§1.3) and that persistent lineages can propagate across Thiès with non-random patterns of movement (§1.4). The spatial kernels identified by these statistical models highlight the underlying local scale of connectivity and movement of lineages within and across years. In comparison, other analyses efforts have focused on population connectivity across longer geographic distances [14, 15, 17]. This work further highlights that combining genetic data and high-resolution metadata (such as GPS and time stamps) with spatial epidemiological modeling has the potential to provide valuable and unique insights into the characteristics and locality of transmission patterns. More practically, these insights could enable better targeting of local interventions by identifying the relative contribution and scale of local transmission versus importation in addition to providing evidence for the role specific neighborhoods play in transmission.

We should emphasize some limitations of this study. The samples were collected opportunistically since 2006 from individuals seeking care at local clinics. Genotyped samples are not uniformly distributed across the malaria seasons, catchment populations, nor directly tied to routine surveillance efforts. In addition to these challenges, approximate Bayesian computation has known short-comings [35], especially in the context of identifying a complete set of summary statistics for model fitting. We have attempted to mitigate the challenge of conducting a retrospective study by using a principled model fitting procedure (Methods) tailored for this dataset (Supplement §3.D), which enables the quantification of uncertainty of these estimates (Fig. 1, Supplement §3.B, 3.C). The detailed epidemiological model provides opportunities to test and validate assumptions. For example, our investigation revealed a mismatch between the transmission dynamics of simulations within season and those of empirical data, highlighting the need for more consistent sampling through the season (§1.2). Moreover, for certain analyses like the local connectivity across neighborhoods, we avoid drawing conclusions from the properties or dynamics of a single lineage, and rather obtain aggregated results across all lineages. We have also used statistical bootstrapping techniques (§1.3) and have posited multiple movement and null models (§1.4) to interrogate our findings.

The 24-SNP barcode also limits the genetic information available for inferring transmission levels and local connectivity. The barcode has been an important tool for investigating malaria parasite genetics [27], but is confined to characterizing identity-by-state and identifying polygenomic infections without resolution of the corresponding parasite subtypes [22]. Recent analyses have leveraged whole genome sequencing data and genetic relatedness features such as identity-by-descent and demonstrated the value for identifying population connectivity [14]. Nonetheless, the 24-SNP barcode can accurately identify clones [22] and produce simple summary statistics that have proven quite powerful for genomic epidemiology.

The work in this article strengthens the evidence for the value of malaria genomic surveillance data for NM-CPs. Genetic features can provide a parsimonious set of indicators to complement routine surveillance, identify local hot-spots of transmission, and monitor the effectiveness of interventions. Epidemiological modeling is poised to be an important tool for these use cases as well as the broader evaluation process for incorporating genomic data into routine malaria surveillance.

## Methods

### 2.1 Data Collection, Sampling, and Barcode Generation

Samples were collected passively from patients reporting to clinics, conditional upon written consent of the subject or a guardian. Subjects were older than 12 years and had onset of acute fever within 24 hours of reporting. Malaria diagnoses were performed with rapid diagnostic tests and microscopic examination. The protocol was approved by the ethical committees of the Senegal Ministry of Health and the Harvard T.H. Chan School of Public Health (Protocol Number 16330). There were 2035 samples genotyped for 24-SNP molecular barcodes [27] from 79 catchment areas in Thiès from 2006-2018.

### 2.2 Epidemiological Model

We constructed a mechanistic model for *Plasmodium falciparum* to infer epidemiological indicators from genetic features (Supplement §2). Seasonal transmission dynamics and outcrossing between parasite haplotypes were simulated together within a simple agent-based framework (Supplement, §2.A, §2.B, §2.C), along with individual strain tracking. Simulations were initialized according to the allele frequencies present in the data from 2006. We approximated the clinic-based collections with passive sampling in the simulations (Supplement §2.D).

### 2.3 Approximate Bayesian Computation and the IMIS Algorithm for Model Calibration

The model was calibrated iteratively via an Incremental Mixture Importance Sampling (IMIS) procedure (Supplement §3). A pseudo-likelihood for the approximate Bayesian computation was constructed using a set of 58 summary statistics of features (Figure 1, panel B, and Supplement §3.A). The pseudo-likelihood was evaluated using a kernel density estimator (KDE), with 100 stochastic replicates per sample (Supplement §3.B).

### 2.4 Genomic feature selection

We leveraged simulations to identify features that are the most informative to predict *R*_0_ (§1.2 and Supplement, §3.C). Spearman correlations and LASSO regression coefficients were calculated with summary statistics as inputs and the *R*_0_ per year as outputs, supplemented by an exploration of the utility of the Akaike information criterion (Supplement Tables S2, S3).

### 2.5 Spatial Analysis of Local Clonal Distributions

Barcode samples were divided into two groups, clones (identical barcodes) and non-clones. The distribution of pairwise distances (derived from GPS coordinates of associated clinic sites) were statistically compared between the two groups (§1.3, Supplement §4A). Bootstrapping was also used to estimate the differences in the mean and median of the resampled distributions (Supplement Fig. S12, S13).

### 2.6 Probabilistic Movement Models for Persistent Strains

We subset the barcodes to persistent monogenomic lineages and then perform a principal component analysis on the pairwise distance and collection time between clonal samples. For each clonal group, we take the component with the largest variance, select those with stronger correlations, excluding statistical outliers, and compute the average (Supplement Fig. S16). Based on the neighborhood locations in Thíes, several probabilistic movement models are generated to compare to empirical calculations. (Supplement §4B).

## Data Availability

Data will be available upon acceptance to peer reviewed journal.

## Acknowledgements

This publication is based on research funded in part by the Bill & Melinda Gates Foundation (BMGF), including modeling and analysis performed by the Institute for Disease Modeling at BMGF. Funding for this work at the Harvard T.H. Chan School of Public Health and the Broad Institute was provided by a BMGF grant to D.F.W. (OPP1156051). We would like to thank the collection team at the SLAP clinic, including Younouss Diedhiou, the Field Lab Coordinator, Dr. Ngayo Sy, the Director, and all the nurses. We would also like to thank the residents of Thiès who have participated in these ongoing studies since 2006. Finally, we thank Amy K. Bei, Sidiya Mbodj, and Fatoumata Dabo for help collecting neighborhood GPS coordinates and harmonizing village names from Wolof for the spatial analysis used in the model.

## References

[1] N Wurtz, et al., Prevalence of molecular markers of Plasmodium falciparum drug resistance in Dakar, Senegal. Malaria Journal 11, 1–10 (2012).

[2] A Dwivedi, et al., Plasmodium falciparum parasite population structure and gene flow associated to anti-malarial drugs resistance in Cambodia. Malaria Journal 15, 319 (2016).

[3] E Talundzic, et al., Next-generation sequencing and bioinformatics protocol for malaria drug resistance marker surveillance. Antimicrobial Agents and Chemotherapy 62 (2018).

[4] WH Organization,, et al., False-negative RDT results and implications of new reports of P. falciparum histidine-rich protein 2/3 gene deletions, (WHO), Technical report (2017).

[5] NMCP, National Malaria Strategic Plan 2014-2020, Abridged version, (The United Republic of Tanzania, Ministry of Health and Social Welfare), Technical report (2014).

[6] USAID, U.S. President’s Malaria Initiative Senegal Malaria Operational Plan FY 2020, (Retrieved from (www.pmi.gov)), Technical report (2020).

[7] MPAC Meeting, Technical consultation on the role of parasite and anopheline genetics in malaria surveillance, (WHO), Technical report (2019).

[8] WH Organization, World malaria report 2020: 20 years of global progress and challenges, (World Health Organization), Technical report (2020).

[9] M Niang, et al., A molecular survey of acute febrile illnesses reveals Plasmodium vivax infections in Kedougou, southeastern Senegal. Malaria Journal 14, 1–8 (2015).

[10] H Herdiana, et al., Malaria risk factor assessment using active and passive surveillance data from Aceh Besar, Indonesia, a low endemic, malaria elimination setting with Plasmodium knowlesi, Plasmodium vivax, and Plasmodium falciparum. Malaria Journal 15, 468 (2016).

[11] RF Daniels, et al., Genetic evidence for imported malaria and local transmission in Richard Toll, Senegal. Malaria Journal 19, 1–8 (2020).

[12] N Obaldia III, et al., Clonal outbreak of Plasmodium falciparum infection in eastern Panama. The Journal of Infectious Diseases 211, 1087–1096 (2015).

[13] FE Saenz, et al., Clonal population expansion in an outbreak of Plasmodium falciparum on the northwest coast of Ecuador. Malaria Journal 14, 1–10 (2015).

[14] AR Taylor, et al., Quantifying connectivity between local Plasmodium falciparum malaria parasite populations using identity by descent. PLoS Genetics 13, 1–20 (2017).

[15] SN Redmond, et al., De Novo Mutations Resolve Disease Transmission Pathways in Clonal Malaria. Molecular Biology and Evolution 35, 1678–1689 (2018).

[16] HH Chang, et al., Mapping imported malaria in Bangladesh using parasite genetic and human mobility data. eLife 8, e43481 (2019).

[17] A Amambua-Ngwa, et al., Long-distance transmission patterns modelled from SNP barcodes of Plasmodium falciparum infections in The Gambia. Scientific Reports 9, 1–9 (2019).

[18] S Tessema, et al., Using parasite genetic and human mobility data to infer local and cross-border malaria connectivity in Southern Africa. eLife 8, e43510 (2019).

[19] I Omedo, et al., Micro-epidemiological structuring of Plasmodium falciparum parasite populations in regions with varying transmission intensities in Africa. Wellcome Open Research 2, 10 (2017).

[20] DE Neafsey, SK Volkman, Malaria genomics in the era of eradication. Cold Spring Harbor Perspectives in Medicine 7, a025544 (2017).

[21] R Dalmat, B Naughton, TS Kwan-Gett, J Slyker, EM Stuckey, Use cases for genetic epidemiology in malaria elimination. Malaria Journal 18, 163 (2019).

[22] RF Daniels, et al., Modeling malaria genomics reveals transmission decline and rebound in Senegal. Proceedings of the National Academy of Sciences of the U.S.A. 112, 7067–7072 (2015).

[23] OJ Watson, et al., Evaluating the Performance of Malaria Genetics for Inferring Changes in Transmission Intensity Using Transmission Modeling. Molecular Biology and Evolution 38, 274–289 (2021).

[24] JA Hendry, D Kwiatkowski, G McVean, Elucidating relationships between P. falciparum prevalence and measures of genetic diversity with a combined genetic-epidemiological model of malaria. bioRxiv (2020).

[25] A Lee, et al., A detailed model of P. falciparum recombination runs modularly on transmission trees to provide new insights on population genetic dynamics (Presentation, ASTMH) (2020).

[26] J Russell, et al., A spatial epidemiological-genetic model to support country program decision-making in malaria control and elimination strategy in Mozambique (ASTMH) (2020).

[27] R Daniels, et al., A general SNP-based molecular barcode for Plasmodium falciparum identification and tracking. Malaria Journal, 223 (2008).

[28] R Daniels, et al., Genetic surveillance detects both clonal and epidemic transmission of malaria following enhanced intervention in Senegal. PloS One 8, e60780 (2013).

[29] W Wong, EA Wenger, DL Hartl, DF Wirth, Modeling the genetic relatedness of Plasmodium falciparum parasites following meiotic recombination and cotransmission. PLoS Computational Biology 14, e1005923 (2018).

[30] KB Gustafson, JL Proctor, Identifying spatio-temporal dynamics of Ebola in Sierra Leone using virus genomes. Journal of the Royal Society Interface 14, 20170583 (2017).

[31] JV Ribado, et al., Linked surveillance and genetic data uncovers programmatically relevant geographic scale of Guinea worm transmission in Chad. medRxiv (2020).

[32] Dmsb Mouzin E, Thior PM, Focus on Senegal Roll Back Malaria: Progress and Impact Series, (WHO), Technical report (2010).

[33] DJ Weiss, et al., Mapping the global prevalence, incidence, and mortality of Plasmodium falciparum, 2000–17: a spatial and temporal modelling study. Lancet 394, 322–331 (2019).

[34] PMI, PRESIDENT’S MALARIA INITIATIVE Senegal Malaria Operational Plan FY 2014 (2014).

[35] K Csilléry, MG Blum, OE Gaggiotti, O Françcois, Approximate Bayesian computation (ABC) in practice. Trends in Ecology and Evolution 25, 410–418 (2010).

