## Supplemental Materials for "Modeling the levels, trends, and connectivity of malaria transmission using genomic data from a health facility in Thiès, Senegal"

September 15, 2021

#### 1 Comparisons with Previous Results

##### 1.1 Piecewise Parameterization Results for $R_0$

We compare calibration results of the genomic epidemiological model with those published in Daniels et al. [1]. In this section we discuss the piece-wise parameterization described below in Section 2.2.3.1. The model itself is outlined in Section 2 and the summary statistics used to fit to data are listed in Section 3.3.1. We find that the best fit triplet $\vec{R_0}$ is $(2.4, 1.8, 2.0)$, as shown in Figure 1. Figure 3 shows a consistent result from [1].

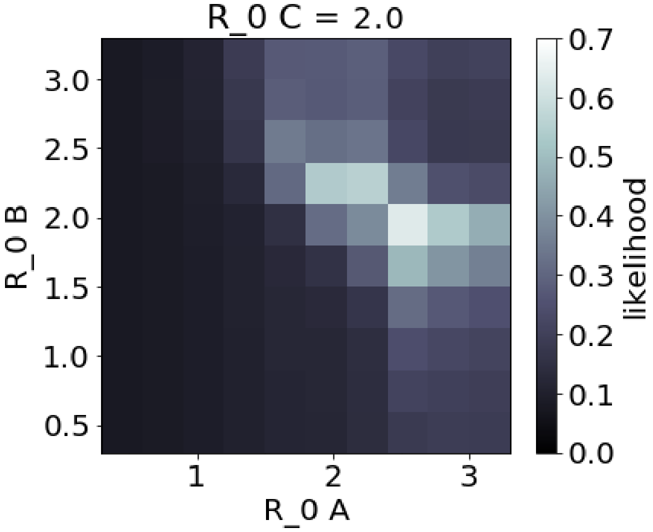


Figure 1: **Likelihood surface for piece-wise parameterization.** Shown above is the log likelihood of the model as a function of $R_{0,A}$ and $R_{0,B}$ when $R_{0,C} = 2.0$. The maximum likelihood triplet is $\vec{R_0} = (2.4, 1.8, 2.0)$.

We note that the model is sensitive to changes in boundary years. That is, summary statistics calculated for the first or last year of data are causally correlated with data or events that happen before or after the duration of data collection. The lack of information pertaining to years beyond the range of the data and corresponding simulations leads to an increase in uncertainty in the results for the first and last years.

Indeed we see that our estimate of $R_{0,C}$, the last parameter in the triplet, (which determines transmission levels in the model from 2010 to 2013) changes in value when we calibrate the model to data collected after 2013 (which was not used in the 2015 model). On the other hand, the values for the first two parameters of the triplet, $R_{0,A}$ and $R_{0,B}$ (which specify transmission from 2006 to 2010), remain consistent since there is no change in the information regarding transmission around 2006 or 2010. When we extend the piece-wise function for data beyond 2013, as detailed below in Section 2, Equation 2.3.1, we get the best fit quadruplet $\vec{R_0} = (2.5, 1.8, 3.0, 1.5)$ (Figure 2).

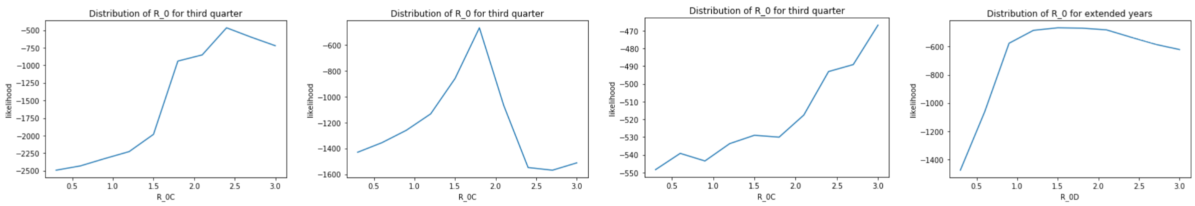


Figure 2: Marginal likelihoods for $R_{0,A}$, $R_{0,B}$, $R_{0,C}$, and $R_{0,D}$. The maximum likelihood set is $\vec{R_0} = (2.5, 1.8, 3.0, 1.5)$. $R_{0,B}$, is lower than the $R_0$ values for years preceding and following, indicating a dip in transmission intensity around 2009.

| | 2006 | 2007 | 2008 | 2009 | 2010 | 2011 | 2012 | 2013 | 2014 | 2015 | 2016 | 2017 | 2018 |
|---|---|---|---|---|---|---|---|---|---|---|---|---|---|
| $E[R_0]$ via KDE | 2.93 | 2.26 | 2.21 | 2.07 | 2.30 | 2.41 | 2.51 | 2.46 | 1.96 | 2.57 | 2.58 | 2.35 | 2.52 |
| KDE 2.5% quantile | 2.58 | 1.78 | 1.70 | 1.66 | 1.66 | 1.62 | 1.96 | 1.73 | 1.56 | 1.75 | 2.04 | 1.57 | 1.63 |
| KDE 97.5% quantile | 3.34 | 2.60 | 2.46 | 2.64 | 2.78 | 3.10 | 3.33 | 3.45 | 3.08 | 3.28 | 3.33 | 3.33 | 3.34 |

Table 1: **Mean $R_0$ and confidence intervals from incremental mixture importance sampling (IMIS) calibration.** The numbers in this table correspond to the positions of the ticks in the second plot of Figure 4.

##### 1.2 Per-Year Calibration Results for $R_0$

For the main article, we fit the data using a per-year model of variation in $R_0$ (Eq. 6). We find that the qualitative trends in $R_0$ are robust to the choice of parameterizations. Both the piecewise and per-year versions show a significant drop and then rebound in transmission from around 2009 to 2012. See Table 1 and Figure 4 for details. The per-year fit shows qualitative agreement with previous results, while also indicating a sharp drop in $R_0$ in 2014. The inferred sharp drop matches the summary statistics; i.e., $f_{unique}$ is almost half that of 2013 (a greater fractional change than almost any other year).

#### 2 The malaria genomic epidemiological model

The genomic epidemiology simulations in this study are carried out by a mechanistic genetic model for malaria transmission (MGMT). It includes two main components: one that governs the mechanism of inheritance of genetic information among successive generations of parasites, and another that controls the epidemiological processes for transmitting parasites to new hosts. In this section, we first summarize the genetic mechanisms and then describe modeling transmission. We then elaborate on the simulation procedure, including the initial conditions and parameters required for simulation. We also describe how we sample the model output.

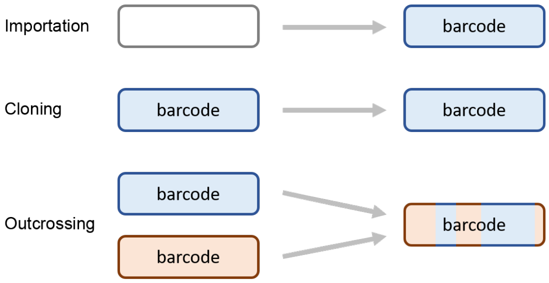


Figure 6: Graphical representations of the three dynamic processes for malaria genomic evolution - importation, clonal propagation, and outcrossing - as outlined in Section 2.1.

##### 2.1 The Genetic Model

Whenever a new infection is generated in the transmission simulation, we assign it a molecular barcode. There are three processes to generating a barcode for the new infection:

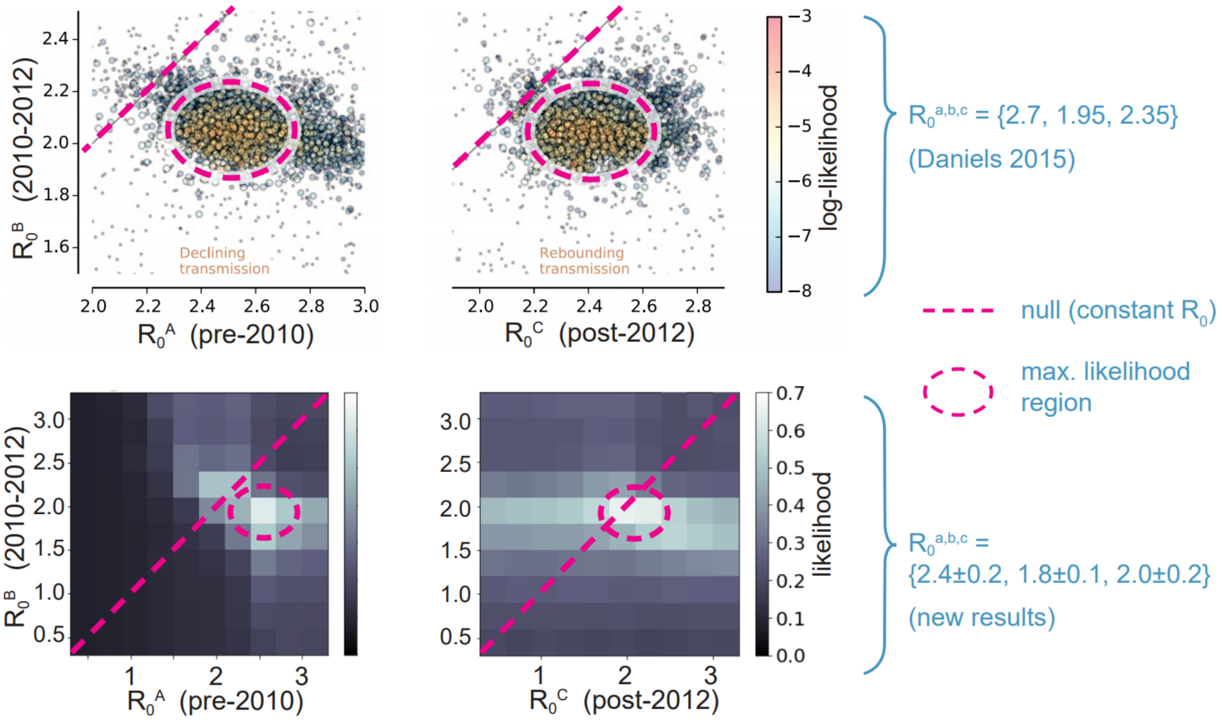


Figure 3: We compare likelihood surfaces from an on-grid parameter sweep of our dynamic model (bottom row) with a scatter plot of likelihoods calculated from an IMIS calibration of the Daniels 2015 model (top row). The top two plots show a cluster of samples in $R_0$ triplet space with a region of high likelihood circled in a dashed pink line. A diagonal dashed pink line indicates the region in parameter space corresponding to a null model with no variation in $R_0$. The bottom two plots show likelihood surfaces corresponding to the same parameter space. The left plot is a slice through the first two parameters $R_{0,A}$ and $R_{0,B}$ at the most likely value of $R_{0,C}$. The right plot is a slice through $R_{0,C}$ and $R_{0,B}$ at the most likely value of $R_{0,A}$. The dashed pink lines once again indicate the high likelihood region and the null model. Both the old model (D15) and the new model (MGMT) reject the null, and have similar qualitative trends for the $R_0$ triplet.

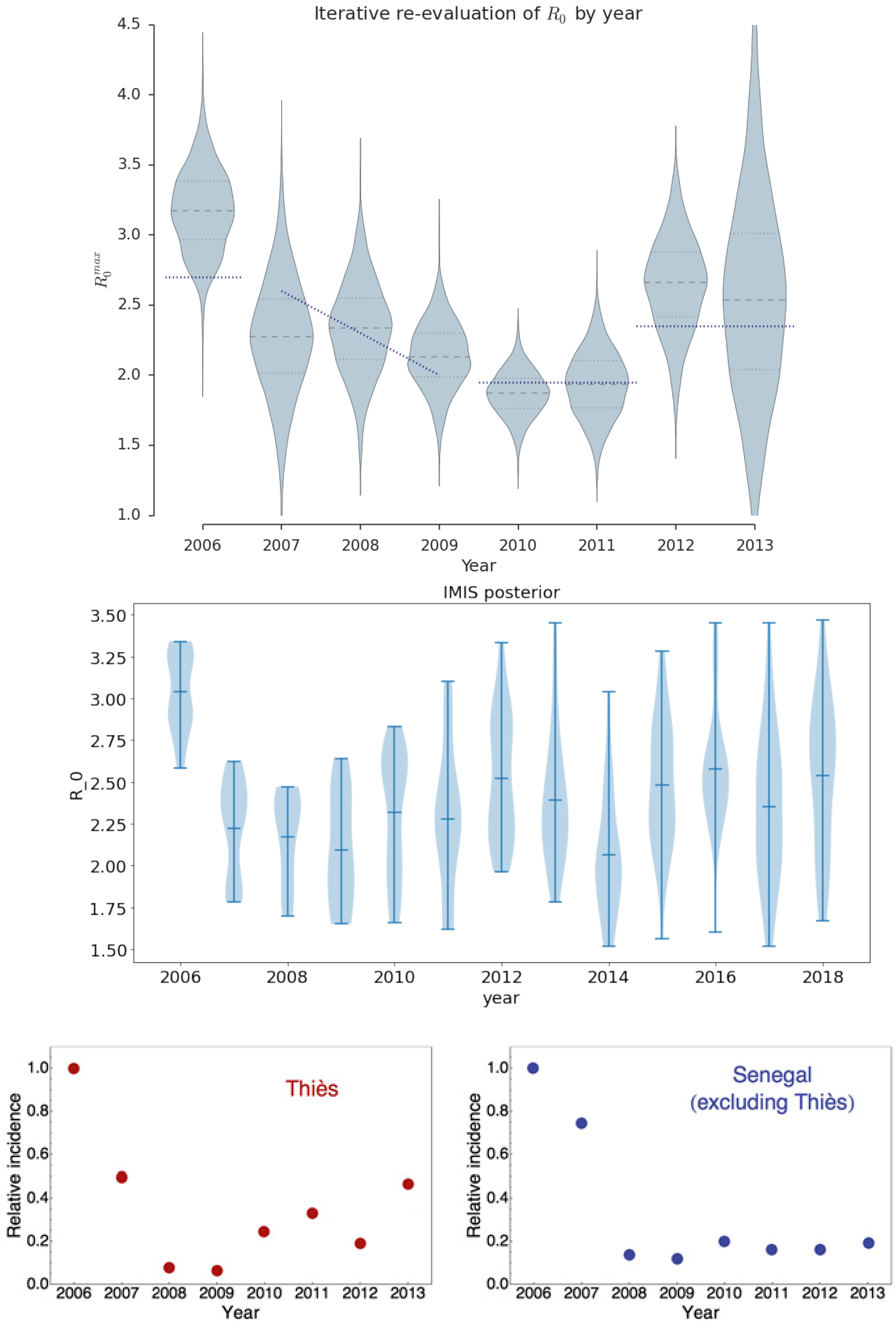


Figure 4: Per-year fits via IMIS, from D15 (top plot) and MGMT (middle plot). Both calibration results show general qualitative agreement, with a decline in transmission starting in 2007 and then a rebound around 2011, which also agrees with incidence data from Thiès (bottom left). Neither model shows correlation with incidence data from other parts of Senegal (bottom right), which suggests that dynamic agent based models may be sensitive to the particular regional transmission dynamics in Thiès. We note that the comparisons with data are only valid for qualitative trends since reported incidence from routine data does not necessarily scale linearly with $R_0$ over broad transmission regimes, and MGMT does not take health-seeking behavior into account. Upcoming data from national surveillance across Senegal will allow us to test the specificity of this modeling framework and whether it can distinguish between dynamics from different regions. For the MGMT calibration, feature selection was used to identify statistics that were more informative for inferring incidence, which may be a reason for its higher correlation with the incidence data from Thiès.

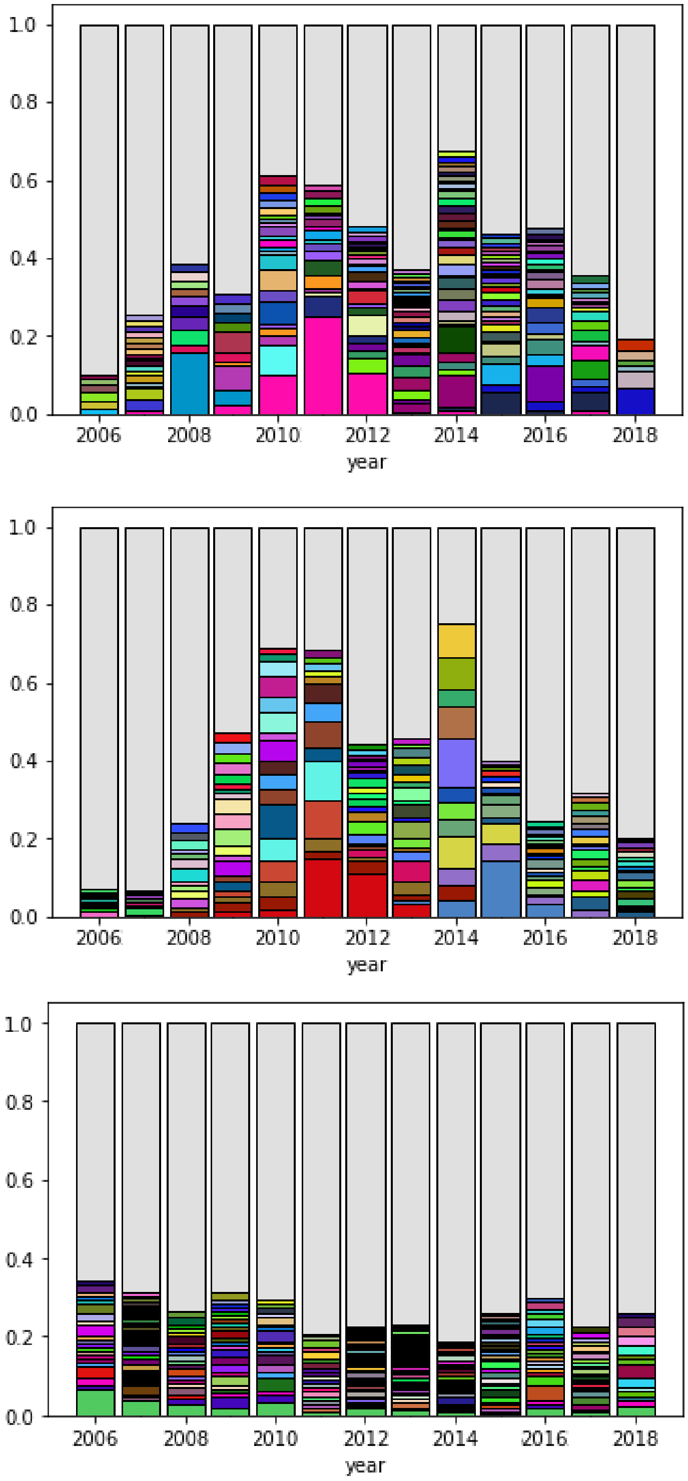


Figure 5: Here we visualize some of the summary statistics used in the calibration. Each repeated strain is given a corresponding color, while unique strains are shown in aggregate as the grey bars. The diagrams show the fraction of unique strains changing per year to year, as well as some persistent strains that exist across many years. The top figure shows the statistics calculated from the Thiès data, the middle figure shows the same for a stochastic simulation using the best fit parameterization, and the bottom for a simulation of a null scenario with no variation in $R_0$. The calibrated model is a better approximation of the data, with notable features such as a reduction in unique strains in 2014 as well as a large clonal expansion of a single strain from 2010 to 2012.

- Generate a new barcode where each of the unlinked 24 SNP positions is an independent random draw weighted by the population allele frequencies (importation).

- Select an existing barcode in the parasite reservoir and pass it to the new infection (clonal propagation).

- Choose two existing barcodes in the active parasite reservoir and perform meiotic recombination between the barcodes to generate one offspring barcode (outcrossing)

This genomic model is adapted from Daniels et al. [1], and details about the outcrossing mechanism and linkage rates can be found in its supplemental materials. In Figure 6, we provide schematic diagrams for each of the above methods for generating barcodes. The simplified model for outcrossing does not simulate the effects of having multiple related haploid offspring.

#### 2.2 Transmission Model

Here, we describe the transmission model. We use an agent-based modeling framework allowing for explicit tracking of individual strains for polygenomic infections. We describe this modeling innovation which is used for the results in the main manuscript (§2.2.1). We also describe the model in Daniels et al. [1] for comparison (§2.2.2).

##### 2.2.1 Agent-based model with explicit strain tracking

The agent-based model keeps track of a list of human hosts among which parasites can be transmitted. Each host can hold up to 10 haplotypes, and whenever a reinfection occurs, we represent this by adding a new haplotype to the current list of haplotypes infecting the host (allowing for duplicates). This new framework allows us to simulate the effects of multiple infections without losing resolution on genetic strains that are propagated in the population. In order to compare the results of these simulations with data from Senegal, whenever an an individual is sampled, the set of haplotypes currently in the infected host are aggregated into a single barcode where SNP positions that have mixed alleles across the haplotypes are called an N. This information is kept in a table called Infection History. If only one haplotype is currently infecting the host, then the monogenomic barcode is recorded as is. The total set of barcodes is kept in the Reservoir where each barcode representation remains un-aggregated so that the genomic information of each haplotype can be preserved. We show the model structure as a schematic diagram in Figure 7 and examples of aggregated barcodes in Figure 8.

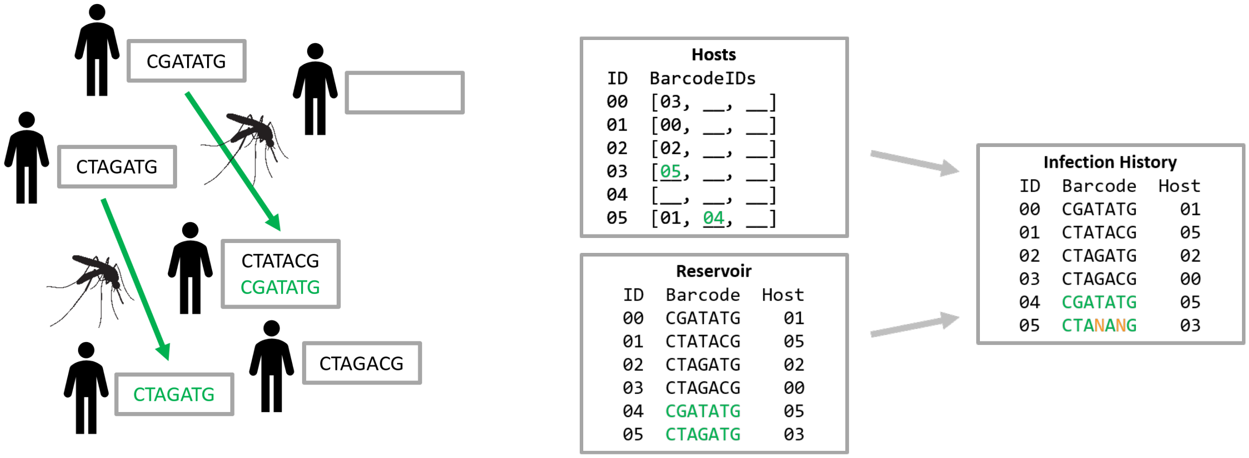


Figure 7: **Model with infection multiplicity.** Each barcode sample is assigned to a human host. Barcodes highlighted in green indicate how the system state is updated with new barcodes after one time step. Note that each host can be infected by multiple strains, but when this information is converted to simulated data, we encode infections as aggregated barcodes.

##### 2.2.2 Previous model from Daniels et al. 2015

Individual haplotypes are not explicitly tracked for polygenomic infection. Instead, polygenomic infections are represented by a single barcode where SNPs with the same allele among all haplotypes in the individual are indicated as is, whereas SNPs with multiple alleles are represented by the letter "N" (Figure 8). When polygenomic infections are transmitted, they are directly transferred as a cotransmission event [1]. This approximation loses information about the diversity of all strains within a single host and potentially within the population. However, the parsimonious barcode model reduces computation time and complexity since there is only one barcode per infection.

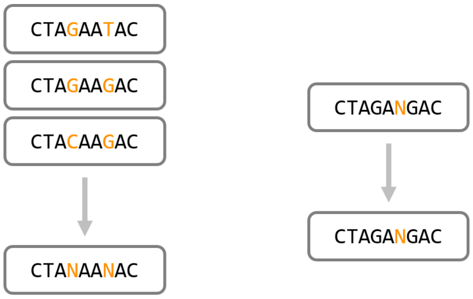


Figure 8: **Polygenomic aggregation.** When a host has a multiple infections, all of the coexistent haplotypes are represented by a single collapsed barcode where "N"s represent the biallelic sites (as shown on the left). In the Daniels 2015 model, polygenomic infections are represented by a single barcode for both the reservoir and infection history.

#### 2.3 Overview of model simulations

In this section, we provide an overview of the algorithm for simulating the model, the initialization of each simulation, and sampling of the reservoir We provide a summary of the simulation software in Algorithm 1.

In detail, a base $R_0$ value is calculated for each time step based on a seasonal function and an overall scaling factor:

$$R_0 = R(t) = R_{scale}(t)\left[\rho + (1-\rho)\cos^2\frac{\pi(t-t_0)}{365}\right] \tag{1}$$

$$R_{adjusted} = R_0\left[1 - \frac{C_{inf}}{5}\right] \tag{2}$$

$$C_{inf} = N_{infected}/N_{hosts} \tag{3}$$

$R_{adjusted}$ is used to calculate the number of actual new infections to generate. We use an adjusted reproductive number to prevent the simulation from generating a runaway outbreak and using up CPU time on a highly unlikely scenario. Since prolonged exponential growth is not something observed in the Senegal data we believe it is reasonable to impose this bound. We then multiply this with $N_{cleared}$ the number of infections cleared at time $t$ to obtain $N_{new} = R_{adjusted}N_{cleared}$. $R_{scale}(t)$ may be a constant, or it may be parameterized to vary over time. We employ a few different methods of formulating $R_{scale}(t)$ in order to calibrate to data, and these are described in Subsection 2.3.1.

$N_{cleared}$ is calculated by the function $f_{count\_expiring\_infections}$ which counts the number of infections that clear at the current time $t$ (Alg. 1 Line 7). In order to approximate an exponential distribution of infection durations, each infection has a fixed probability of clearing at any given time $t$ .

Next we generate a set of new infections as specified by $N_{new}$. In order to determine the mechanism of transmission, we calculate the probability of clonal transmission based on the average COI of the reservoir, and via random draw determine whether or not to simulate a clonal transmission. Based on Poisson statistics we define this probability to be:

$$P_{clonal} = 1 - P_{outcross} = 1 - \sum_{k>=2} \text{Poisson}(k, \lambda = C_{inf}) \tag{4}$$

$$= e^{-C_{inf}} + c_{inf}e^{-C_{inf}} \tag{5}$$

If a clonal transmission is triggered, we use $g_{clone}()$ to randomly select a barcode from the existing reservoir $I_R$, and then use $f_{append}()$ to add the selected barcode to a list of new infections $I_t$ (Alg. 1 Line 13). If an outcrossing event is triggered, we instead use $g_{meiosis}()$ to randomly select two barcodes from $I_R$ and generate a meiotic offspring, as detailed in [1], and append it to $I_t$ (Line 15).

**Algorithm 1** Stochastic simulation procedure for mechanistic genomic model for malaria transmission

**Input:** Specify the following: time step size $\delta t$, start and end dates for simulation $t_0$ and $t_{end}$, importation rate $p_i$, daily infection clear rate $p_K$, maximum allowed number of infections $l_{max}$

1: **procedure** BarcodeModel
2: Initialize calibration parameters $\{R_{0,i}\}$ (Eq. 6)
3: Initialize state variables: $I_R, I_H, t$ ▷ reservoir, infection history, and start time
4:
5: **while** ($t$ ¡= $t_{end}$) and (length($I_H$) ¡ $l_{max}$) **do**
▷ Simulate transmission.
6: $R_{adjusted} \leftarrow R(t, \{R_{0,i}\})$ ▷ We are interested in calibrating $R(t)$.
7: $N_{cleared} \leftarrow f_{count\_expiring\_infections}(I_R, p_K)$ ▷ Count infections to be cleared at time $t$ (see §2.3)
8: $N_{new} \leftarrow R_{adjusted} N_{cleared}$
9: $I_t \leftarrow \varnothing$ ▷ Initialize temporary list of new infections $I_t$ for current time step $t$.
10:
11: **for** each instance $j \in \{1, ..., N_{new}\}$ **do**
▷ Simulate genetics.
12: **if** $r_{random}() < P_{clonal}$ **then** ▷ $P_{clonal}$ defined in Eq. 4
13: $I_t \leftarrow f_{append}(I_t, g_{clone}(I_R))$ ▷ Pick random strain from $I_R$ and append to $I_t$.
14: **else**
15: $I_t \leftarrow f_{append}(I_t, g_{meiosis}(I_R))$ ▷ Generate new infection via outcrossing and append to $I_t$.
16: **end if**
17: **end for**
18:
19: **if** $r_{random}() < p_i$ **then**
20: $I_t \leftarrow f_{append}(I_t, g_{import}())$
21: **end if**
22:
23: $I_H \leftarrow f_{append}(I_H, f_{resolve\_polygenomic}(I_t))$ ▷ Add $I_t$ to history, applying any observation model as needed.
24: $I_R \leftarrow f_{append}(I_R, I_t)$ ▷ Add new infections to reservoir.
25: $I_R \leftarrow f_{remove\_dead}(I_R)$ ▷ Remove cleared infections from reservoir.
26: **end while**
27: **end procedure**

Independently of the new infections created by the local reproductive number, we use $g_{import}()$ to generate new imported infections at a rate specified by $p_i$ (Alg. 1 Line 20). $g_{import}()$ simulates new barcodes by randomly assigning alleles for all 24 SNP positions according to the empirical population allele frequencies.

At the end of each time step we delete infections from the reservoir that were marked for expiration (i.e. those counted by $N_{cleared}$) and add newly generated infections to the reservoir and their corresponding observations to the infection history. As discussed in Sections 2.2.1 and 2.2.2, the way infections are recorded in the reservoir and infection history is dependent on the particular per-host model.

After repeating the above for every time step, this results in a history of infections at the end of the simulation.

##### 2.3.1 Model Parameters, Initialization, and Calibration

The dynamic model has a large number of epidemiological input parameters than can be adjusted, but we only vary $R_0$ during calibration since we are interested in linking genetics to transmission intensity. We fix the remaining parameters based on empirical data. For example, the seasonal modulation in vector feeding rates is known to correlate with rainfall, so we pick suitable values for $t_0$ and $\rho$ (Eq. 1) that have been informed by independent data.

We parameterize $R_0$ as a function of time by assigning $R_{scale}$ a constant value for each year:

$$R_{scale}(t) = R_{0,i} \quad \text{for } t_{i-1} < t < t_i \tag{6}$$

We use this parameterization for the iterative calibration method described in Section 3.3.4 and to generate the plots in Figure 4.

For comparisons with Daniels 2015 [1], we use the same piece-wise function from [1], i.e.:

$$R_{scale}(t) = \begin{cases} R_{0,A}, & t \leq t_1 \\ R_{0,A} + (R_{0,B} - R_{0,A})\frac{t-t_1}{t_2-t_1}, & t_1 < t \leq t_2 \\ R_{0,B}, & t_2 < t \leq t_3 \\ R_{0,C}, & t > t_3 \end{cases}$$

where $t_1 = 2007$, $t_2 = 2010$, and $t_3 = 2012$. For data beyond 2013, we let $t_4 = 2014$ and extend the formula:

$$R_{scale}(t) = \begin{cases} R_{0,A}, & t \leq t_1 \\ R_{0,A} + (R_{0,B} - R_{0,A})\frac{t-t_1}{t_2-t_1}, & t_1 < t \leq t_2 \\ R_{0,B}, & t_2 < t \leq t_3 \\ R_{0,C}, & t_3 < t \leq t_4 \\ R_{0,D}, & t > t_4 \end{cases}$$

We define $\vec{R_0}$ to be the list of values $\{R_{0,i}\}$. For the piece-wise functions, we let $\vec{R_0}$ be the triplet or quadruplet of values, $\{R_{0,A}, R_{0,B}, R_{0,C}\}$ or $\{R_{0,A}, R_{0,B}, R_{0,C}, R_{0,D}\}$. For either parameterization, there are discontinuities at the transition between one year to the next. Nevertheless, we see that the discontinuities do not have a significant effect on the simulation dynamics since they occur during the dry season.

The remaining model parameters are fixed. These include:

- Per-day probability of importing a strain from outside the local reservoir, $p_i = .01$ per day
- Per-day probability of any given infection clearing (implies exponential infection durations), $p_K = 0.022$ per day
- Number of human hosts in local population, $N_{hosts} = 1350$
- Initial number of infections in the local reservoir, $N_{infected}(t_0) = 1200$
- Initial fraction of infections that are unique, $f_{unique,0} = 0.9$

We initialize our simulations based on the conditions seen in the Thiès molecular barcode data. We set the diversity of the initial population to match that of the data by using the same allele frequencies, polygenomic fraction $f_{poly}$, and unique fraction $f_{unique}$. Definitions of the latter two statistics are available in Model Evaluation, Section 3.3.1.

##### 2.3.2 Comparison with Epidemiological Model from Daniels et al.

There are important differences between the epidemiological model and fitting procedure in this paper with the Daniels et al. [1]:

- MGMT is an agent based model that keeps track of human hosts and infections separately allowing granular information about multiple infections per host.
- The inclusion of human hosts allows for future model development such as geographic segmentation of human hosts.
- MGMT has options to sample from the entirety of each year, just the rainy season, or a heterogeneous sampling based on the frequency of samples in the real data.
- MGMT has different algorithmic details accounting for the new functionality.*
- MGMT has the option to evaluate likelihoods using kernel densities vs. solely synthetic likelihoods.
- MGMT calculates uncertainties for the synthetic likelihoods using different algorithms.*

Nearly all items above can be parameterized in the model implementation to require MGMT to behave identically to the original model, except for those indicated by an asterisk (*). This results in two key differences that cannot be reverted: the order of operations, and the method for calculating uncertainties. Despite these algorithmic differences, we find quantitative similarity to the results generated by Daniels et al. which provides a foundation for increasing the model complexity and biological fidelity. There are, however, small quantitative differences in the model fits based on the fitting procedure, i.e., a sequential fitting of the reproductive number vs. fitting all of the parameters at once.

#### 2.4 Sampling Models

As noted in Section 2.3.2 we explored three different methods for sampling the final history of infections:

1. **Uniform sampling:** Draw samples from the entire simulation with uniform probability per infection. The total number of samples matches the total number of barcodes from the Thiès data; however, the relative number of samples per year are proportional to the simulation, not the data.

2. **Uniform semiannual sampling:** For each year, draw samples from just the rainy season (approximately August to January) with uniform probability per infection for any given year. The number of simulated samples from each year matches the number in the data.

3. **Empirical sampling:** Randomly draw simulation samples from each month to match the number of barcodes in the data for the corresponding month. The number of samples in each month and year are now similar to those for the real data. One consequence of this is that all samples are also drawn from the rainy season.

A drawback of the third method is that the peak transmission month for any given year in a simulation may be different from the peak transmission month in the data. This can lead to biases in the sampling, and for certain stochastic realizations, some months of the simulation may even have insufficient infections from which to take samples. We therefore primarily use the second method (uniform semiannual sampling) for the results in this paper.

### 3 Model Evaluation

The model is evaluated using approximate Bayesian computation which uses summary statistics to compare simulation outputs to real data. Since the simulations are stochastic, we simulate the model many times to quantify the uncertainty in the model output. The model is calibrated to data using incremental mixture importance sampling (IMIS), as detailed in Subsection 3.4 below. In this section we first outline the summary statistics used for making these comparisons, and then we lay out a formal mathematical framework for calculating likelihoods and comparing models based on the above principles.

#### 3.1 Summary Statistics

The following is a list of summary statistics used for model calibration, which have been chosen for their link to transmission dynamics, based on either correlations with model outputs or references in literature [1]:

- $f_{P,y}$, the fraction of barcodes that are polygenomic for a given year $y$, for each year of data

- $f_{U,y}$, the fraction of barcodes in a given year that are unique across all samples (including polygenomic samples), for each year of data

- $n_{2,y}$, the number of barcodes that are repeated twice within a given year, for each year of data

- $n_{m,y}$, the number of barcodes that are repeated more than twice within a given year, for each year

- $n_{p,x}$, the number of barcodes that persist for $x$ years (for $x = 2, 3, 4$ and $x > 5$)

- $n_a$, the number of barcodes persisting for multiple years that first appear during or before the first year of samples

- $n_d$, the number of barcodes persisting for multiple years that are observed in the final year of samples

In total this results in $4 * n_{year} + 6$ statistics. For the initial versions of the model using data from 2006 to 2013, this equals 38 statistics, and for the newer versions utilizing data until 2018, it equals 58 statistics. Figure 5 shows a visualization of these summary statistics for different scenarios. Please see section 3.3 for a discussion about correlations resulting from the inherent causal nature of the simulations.

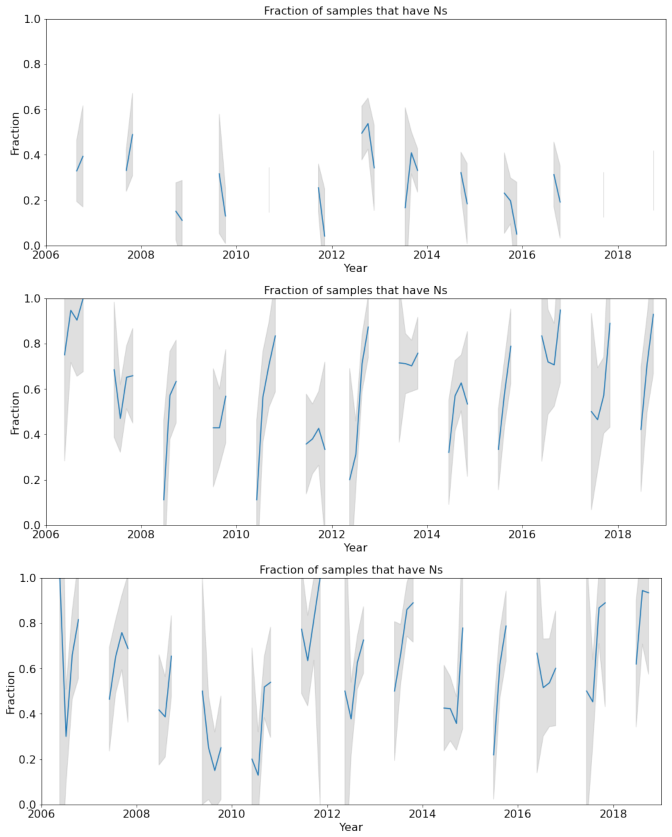


Figure 9: **Temporal trends in polygenomic fractions.** We show the temporal variation in the polygenomic fraction, aggregated into two-month bins. From top to bottom, the plots show the polygenomic fraction calculated from data, from a replicate of the maximum-likelihood parameterization, and from a replicate of the fifth most likely parameterization. The gray bands indicate the $1\sigma$ uncertainty interval. Although individual years show large variation, the general trend across years consistently shows a decrease in the polygenomic fraction from 2006 to 2010, a rebound around 2012, and a drop again around 2014.

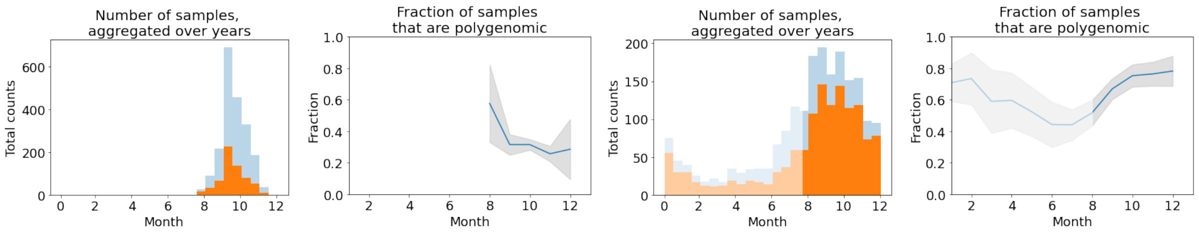


Figure 10: **Aggregated seasonal trends in the polygenomic fraction.** Temporal variation in the polygenomic fraction (as shown in Figure 9) aggregated over years to show trend over an average season. The leftmost plot shows the total number of samples (gray) and the number of polygenomic samples (orange) for data, sorted into two-week bins and aggregated across all years. The next plot shows the same data but expressed as the fraction of samples per half-year bin that are polygenomic (blue line). The gray bands indicate the $1\sigma$ uncertainty interval. The right two figures show the same pair of plots but for simulations.

##### 3.1.1 Polygenomic fraction within malaria seasons

As seen in Model Evaluation Subsection 3.3, the fraction of infections that are polygenomic at a given time $t$ is strongly correlated with transmission intensity. Here, we consider a sample to be polygenomic if there is at least one mixed allele labeled N. All samples with timestamps are included in this analysis. We observe that the polygenomic fraction changes over the course of a malaria season. In Figure 9, we see that the model is able to capture a distinctively rebounding trend from 2010 to 2013 observed in the data, although the variation within a single year is more uncertain. The year-to-year levels of polygenomic fraction tends to be higher overall for the model simulations. There is significant variation between realizations of the model and between seasons in the data. Even aggregating the data across all years, the trends differ between the model simulation and the data within a transmission season (Fig. 10). The empirical data indicates that the polygenomic fraction may be high at the beginning of sample collection and trend lower over a season. Comparing the model output to the data is challenging based on several factors: the transmission season starts at slightly different times each year, the sample collection period is not uniform over a season across all years of study, and the measurement uncertainty grows as there are fewer samples by month or week. An important future research direction is to understand this discrepancy; this will require both a deeper understanding of the epidemiology of malaria transmission within and between seasons as well as a more systematic collection of samples through a season.

#### 3.2 Likelihood Evaluation with ABC

In order to evaluate likelihoods, we use a version of approximate Bayesian computation with a kernel density estimator (KDE). In this section we provide a concise overview of the basic principles of ABC as they pertain to our use case. The main goal of this method is to be able to calculate the likelihood $L(\vec{S}_{obs} \mid M)$ accurately when it is either unknown or does not have a simple analytic form. This is critical for the barcode model since its likelihood does not have a tractable analytic expression and since many of its summary statistics are highly non-Gaussian. We assume that the true distribution of $L(\vec{S}_{obs} \mid M)$ is sampled by the stochastic replicates $\{\vec{S}'_i\}$ of the model parameterization $M$. Then, in the limit of having a sufficiently large number of replicates, $(i = 1, 2, ...N_{sims})$, the density of $\{\vec{S}'_i\}$ will be high enough that the likelihood of any given observed outcome $\vec{S}_{obs}$ is simply the number of replicates within an arbitrarily small neighborhood of $\vec{S}_{obs}$,

$$L(\vec{S}_{obs} \mid M) = \lim_{\epsilon \to 0, N_{sims} \to \infty} L_{count}(\vec{S}_{obs} \mid M, N_{sims}, c_{seed}), \tag{7}$$

$$L_{count}(\vec{S}_{obs} \mid M, N_{sims}, c_{seed}) \equiv \frac{Count(\{\vec{S}'_i \mid M, g(\vec{S}'_i, \vec{S}_{obs}) < \epsilon\})}{Count(\{\vec{S}'_i \mid M\}) \int_{g(\vec{S}, \vec{S}_{obs}) < \epsilon} dV} \equiv \frac{\sum_{i \mid M, g(\vec{S}'_i, \vec{S}_{obs}) < \epsilon} 1}{N_{sims} \int_{g(\vec{S}, \vec{S}_{obs}) < \epsilon} dV}, \tag{8}$$

where $g$ is some distance metric and $dV$ is the differential volume corresponding to the integral over $\vec{S}$. The advantage of this so-called rejection algorithm is that it holds for any finite volume enclosed by an orientable surface $g(\vec{S}, \vec{S}_{obs}) = \epsilon$.

However, this brings to light a significant drawback: in order to have a non-noisy and accurate estimator, $N_{sims}$ must be very large, which is computationally expensive.

##### 3.2.1 Using a KDE to Calculate Likelihoods

In order to approximate $L_{count}(\vec{S}_{obs} \mid M, N_{sims}, c_{seed})$ at low $N_{sims}$, we use a kernel to estimate the frequency of samples:

$$L_{count}(\vec{S}_{obs} \mid M, N_{sims}, c_{seed}) \sim \frac{1}{N_{sims}} \sum_{\vec{S}'_i \mid M} k(\vec{S}'_i, \vec{S}_{obs}), \tag{9}$$

$$\int k(\vec{S}_i, \vec{S}) dS = 1, \tag{10}$$

$$k(\vec{S}_i, \vec{S}) \equiv k(\vec{S} - \vec{S}_i). \tag{11}$$

For the current work we find a normally distributed kernel $k = \exp\left[(\vec{S} - \vec{S}_i)^T \Sigma^{-1} (\vec{S} - \vec{S}_i)\right]$ to be sufficiently accurate for estimating likelihoods. Although $k$ is better at describing the distributions of count statistics such as $n_2$ than those of fractional statistics like $f_P$, the resulting likelihood estimates are comparable to those calculated using a rejection algorithm (Eq. 8). The covariance matrix $\Sigma$ is chosen to be diagonal and the diagonal values $\sigma^2_{kernel,j}$ are determined by two constraints: for each statistic $s_j$, (1) $\sigma^2_{kernel,j}$ must be less than or equal to $\sigma^2_{replicates,j}$, the variance across all replicates $i$ of all model parameterizations $M$, and if possible, (2) $\sigma^2_{kernel,j}$ should be greater than $\sigma^2_{mean,j} = \max(\{\sigma^2_{mean,j,M} \mid Q(M)\})$, where $\sigma^2_{mean,j,M}$ is the variance in the mean of a given parameterization $M$ and $Q(M)$ designates models included in the likelihood comparisons. We find that the qualitative trend in $R_0$ for different model realizations is robust to most choices of covariance matrices within these constraints. However, there can be minor quantitative differences, so the results in this manuscript use the following definition for consistency:

$$\sigma^2_{kernel,j} = \min(\sigma^2_{mean,j} + \sigma^2_{0,j}, \quad \sigma^2_{replicates,j}) \tag{12}$$

$$\sigma^2_{0,j} = E[s_j] \quad \text{if } s_j \in \{n_2, n_{multi}, n_{persist,x}, n_{appear}, n_{disappear}\} \tag{13}$$

$$\sigma^2_{0,j} = E[s_j](1 - E[s_j]) \quad \text{if } s_j \in \{f_{poly}, f_{unique}\} \tag{14}$$

where $\sigma^2_{0,j}$ is the theoretical variance of a statistic $s_j$ with either a Poisson (Eq.13) or binomial distribution (Eq.14).

##### 3.2.2 Synthetic Likelihoods

We also evaluate the model using synthetic likelihoods to compare to previous work [1]. In the limit that summary statistics have a roughly Gaussian distribution, we can make the simplifying assumption that their distributions can be fully specified by calculating their means and covariances. This method is faster than using KDE, and has better noise properties when there are a low number of samples, making it an attractive alternative for quick analyses.

It turns out that, despite its low accuracy for non-Normal distributions, the synthetic likelihoods recover most of the *qualitative* variation in $R_0$ over the course of the simulations. Both synthetic and KDE likelihoods show a decline and then rebound in transmission around 2012. The concordance is largely due to the tightly bound distributions for most of the informative statistics, such as $f_P$ and $f_U$. Nevertheless, for precision calibrations we opt to use KDE likelihoods.

#### 3.3 Identifying informative genetic features for transmission inference

We use the summary statistics and model fitting procedure to identify and rank the most important genetic features for identifying transmission values. ABC has known challenges relative to model mis-specification and not including a complete set of summary statistics. Here, we have a set of 58 summary statistics, many of which are co-linear and correlated. We try to identify a minimal number of informative features for the fitting procedure, thereby reducing the computational complexity and helping characterize the most informative features associated with transmission.

We use various data-driven methods to select features that exhibit stronger correlation with $R_0$ or are more informative for predicting $R_0$. Spearman coefficients between all $R_0$ parameters and all genetic features indicate that the polygenomic fraction $f_P$ is consistently correlated with $R_0$ across all years of data, followed by $f_U$ and $n_2$ (Table 2). All three features are found to be secondary lagging indicators for some years. We also report the decrease in the Akaike information criterion (AIC) when excluding a particular statistic, which suggests that $f_U$ and $f_P$ are better able to constrain the model, but this result is not as reliable since it is dependent on the likelihood estimation.

Least-absolute-shrinkage-and-selection-operator (LASSO) regression is a method for identifying informative features. We find that the regression identifies $f_P$ as the most predictive covariate for every year. This consistency suggests that the selection of $f_P$ is not a chance result from multiple co-linear candidates, but rather an indication that $f_P$ captures critical information about the model dynamics. Statistics have different dynamic ranges, and the relative strength of correlation with other parameters like $f_U$ and $n_2$ changes as $R_0$ fluctuates, suggesting that different features may be more informative for different transmission regimes. The regression also occasionally identifies $f_P$ from the previous year or $f_U$ from the same year as additional informative covariates. This suggests that features can also be leading indicators for certain metrics, and it corroborates earlier work that clonality is an important signal in a low transmission setting like Thiès. The following are a few key takeaways:

1. $f_P$ has the strongest correlation with the input parameters $R_0$, followed by $f_U$ and $n_2$
2. Statistics calculated for a particular year are most correlated with the $R_0$ parameter for the same year.
3. Statistics calculated for a given year $s_{i,y}$ can be dependent on the $R_0$ parameter for previous years $y-1$. This confirms the causal dynamics of the model.

#### 3.4 IMIS

Importance sampling allows us to approach model evaluation in a natural Bayesian manner. Samples for evaluation $\{\vec{\phi}_i\}$ are drawn from a distribution defined by a probability density function (PDF) $q$ such that $P_q(\vec{\Phi} = \vec{\phi}) = q(\vec{\phi})$, and then they are reweighted by a factor $c_i = L(\vec{s} \mid \vec{\phi}_i)\Pi(\vec{\phi}_i)/q(\vec{\phi}_i)$ that produces a new weighted distribution

| | $R_{0,y}$ | | | | | | AIC |
|---|---|---|---|---|---|---|---|
| | $y = 2006$ | 2007 | 2008 | 2012 | 2013 | 2014 | |
| $f_{U,2006}$ | 0.67 | 0.10 | 0.03 | 0.00 | −0.07 | 0.07 | 40.35 |
| $f_{U,2007}$ | 0.24 | 0.85 | 0.08 | −0.00 | −0.08 | −0.03 | 40.34 |
| $f_{U,2008}$ | 0.02 | 0.68 | 0.66 | 0.03 | −0.03 | 0.02 | 40.28 |
| $f_{U,2012}$ | 0.00 | −0.03 | 0.04 | 0.59 | −0.04 | −0.04 | 40.27 |
| $f_{U,2013}$ | −0.04 | −0.07 | 0.01 | 0.30 | 0.76 | 0.04 | 40.27 |
| $f_{U,2014}$ | 0.02 | −0.12 | 0.01 | 0.06 | 0.49 | 0.72 | 40.03 |
| $f_{P,2006}$ | 0.93 | 0.00 | −0.03 | 0.00 | −0.04 | 0.07 | 39.52 |
| $f_{P,2007}$ | 0.33 | 0.87 | 0.07 | 0.02 | −0.07 | −0.02 | 40.17 |
| $f_{P,2008}$ | 0.09 | 0.62 | 0.74 | 0.01 | −0.01 | 0.03 | 39.82 |
| $f_{P,2012}$ | 0.10 | 0.05 | 0.07 | 0.74 | −0.05 | 0.01 | 40.23 |
| $f_{P,2013}$ | 0.08 | −0.01 | 0.04 | 0.34 | 0.87 | 0.02 | 40.05 |
| $f_{P,2014}$ | 0.09 | −0.04 | 0.06 | 0.09 | 0.48 | 0.83 | 40.19 |
| $n_{2,2006}$ | −0.60 | −0.07 | −0.04 | −0.03 | 0.02 | −0.08 | 37.96 |
| $n_{2,2007}$ | −0.24 | −0.81 | −0.10 | 0.02 | 0.07 | 0.02 | 36.28 |
| $n_{2,2008}$ | −0.00 | −0.53 | −0.64 | −0.04 | 0.02 | −0.03 | 36.39 |
| $n_{2,2012}$ | −0.02 | 0.03 | −0.05 | −0.66 | 0.02 | 0.00 | 34.90 |
| $n_{2,2013}$ | 0.03 | 0.08 | −0.03 | −0.29 | −0.78 | −0.03 | 36.60 |
| $n_{2,2014}$ | −0.01 | 0.08 | 0.01 | −0.01 | −0.40 | −0.75 | 34.43 |
| $n_{m,2006}$ | −0.39 | −0.02 | 0.00 | −0.03 | 0.03 | −0.07 | 38.05 |
| $n_{m,2007}$ | −0.22 | −0.68 | −0.04 | −0.04 | 0.08 | 0.02 | 37.86 |
| $n_{m,2008}$ | −0.02 | −0.64 | −0.56 | −0.01 | 0.03 | −0.03 | 37.73 |
| $n_{m,2012}$ | −0.01 | 0.01 | −0.04 | −0.50 | 0.05 | 0.02 | 34.41 |
| $n_{m,2013}$ | 0.04 | 0.06 | 0.02 | −0.23 | −0.71 | −0.06 | 35.41 |
| $n_{m,2014}$ | −0.02 | 0.10 | −0.03 | −0.06 | −0.51 | −0.64 | 36.21 |
| $n_{p,2}$ | −0.14 | −0.26 | −0.23 | −0.15 | −0.06 | −0.28 | 34.81 |
| $n_{p,3}$ | −0.13 | −0.21 | −0.21 | −0.07 | 0.08 | −0.00 | 35.94 |
| $n_{p,4}$ | −0.14 | −0.16 | −0.18 | 0.03 | −0.02 | 0.04 | 37.24 |
| $n_{p,5}$ | −0.14 | −0.11 | −0.08 | 0.03 | 0.12 | 0.09 | 38.13 |
| $n_a$ | −0.14 | −0.23 | −0.24 | −0.30 | −0.34 | −0.32 | 34.66 |
| $n_d$ | −0.16 | −0.23 | −0.25 | −0.30 | −0.34 | −0.33 | 34.30 |

Table 2: Spearman correlations between summary statistics and $R_0$. For brevity we only show the years 2006, 2007, 2008, 2012, 2013, and 2014. Across all years we see that $f_P$ is most correlated with $R_0$ for any given year, followed by $f_U$ and $n_2$. We also see that the statistics for a given year are strongly correlated with the $R_0$ for the same year, however, there can be a less pronounced lagging effect where $R_0$ can still have an impact on future years. We also show the decrease in the Akaike Information Criterion (AIC) when excluding the statistic given in particular row. I.e., the greater the difference, the more informative the statistic. One flaw in this metric is that statistics that provide poor fits to data are also considered informative if they cause a large difference in the likelihood.

described by the posterior $P(\vec{\Phi} = \vec{\phi} \mid \vec{s}) = L(\vec{s} \mid \vec{\phi})\Pi(\vec{\phi})$. We can get a reweighted estimate of any statistic or quantity $f$ via the weighted expected value:

$$E[f] = \frac{\sum_{i=1}^{N} f(\vec{\phi}_i)L(\vec{s} \mid \vec{\phi}_i)\Pi(\vec{\phi}_i)/q(\vec{\phi}_i)}{\sum_{i=1}^{N} L(\vec{s} \mid \vec{\phi}_i)\Pi(\vec{\phi}_i)/q(\vec{\phi}_i)} \tag{15}$$

where $N$ is the total number of samples.

The incremental aspect of IMIS comes from iteratively refining the PDF $q$ to get closer to the posterior. As long as each iteration of the sampling distribution has a closed-form expression, we can produce a mixed distribution defined by the new PDF

$$q'(\vec{\phi}) = \frac{q_1(\vec{\phi}) + q_2(\vec{\phi}) + \cdots + q_l(\vec{\phi})}{l} \tag{16}$$

where $l$ is the number of iterations and $q'(\vec{\phi})$ describes the distribution we would be sampling from if we were to combine all the samples from all iterations into one set, assuming each iteration has the same number of samples. I.e.,

$$\{\vec{\phi}_i\}_{q'} = \{\vec{\phi}_i\}_{q_1} \cup \{\vec{\phi}_i\}_{q_2} \cup \cdots \cup \{\vec{\phi}_i\}_{q_l} \tag{17}$$

$$\{\vec{\phi}_i\}_{q_k} \equiv \{\vec{\phi}_i \mid i \in S_k\} \tag{18}$$

where $S_k$ is the subset of indices $i$ of samples $\vec{\phi}_i$ drawn from $q_k$.

| | $R_{0,y}$ | | | | | | | | | | | | |
|---|---|---|---|---|---|---|---|---|---|---|---|---|---|
| | $y = 2006$ | 2007 | 2008 | 2009 | 2010 | 2011 | 2012 | 2013 | 2014 | 2015 | 2016 | 2017 | 2018 |
| $f_{U,2006}$ | −0.00 | −0.00 | 0.00 | −0.00 | −0.00 | −0.00 | −0.00 | −0.00 | 0.00 | −0.00 | −0.00 | −0.00 | −0.00 |
| $f_{U,2007}$ | −0.00 | 0.00 | −0.00 | 0.00 | −0.00 | −0.00 | 0.00 | −0.00 | −0.00 | −0.00 | −0.00 | 0.00 | 0.00 |
| $f_{U,2008}$ | −0.00 | 0.00 | 0.00 | −0.00 | 0.00 | 0.00 | 0.00 | 0.00 | −0.00 | 0.00 | −0.00 | 0.00 | 0.00 |
| $f_{U,2009}$ | 0.00 | −0.00 | 0.00 | 0.00 | −0.19 | 0.00 | 0.00 | −0.00 | 0.00 | −0.00 | −0.00 | 0.00 | 0.00 |
| $f_{U,2010}$ | 0.00 | 0.00 | −0.00 | 0.18 | −0.01 | −0.00 | 0.09 | −0.10 | −0.00 | 0.00 | 0.00 | −0.00 | −0.00 |
| $f_{U,2011}$ | −0.00 | 0.00 | 0.00 | −0.00 | 0.00 | −0.38 | −0.00 | 0.00 | −0.04 | 0.00 | 0.00 | −0.00 | −0.00 |
| $f_{U,2012}$ | −0.00 | 0.00 | −0.00 | 0.00 | −0.00 | 0.00 | −1.11 | 0.00 | 0.00 | −0.00 | 0.00 | 0.00 | 0.00 |
| $f_{U,2013}$ | −0.00 | 0.00 | −0.00 | 0.00 | −0.00 | 0.00 | 0.00 | −0.88 | 0.00 | 0.00 | −0.00 | 0.00 | 0.00 |
| $f_{U,2014}$ | 0.00 | −0.00 | 0.00 | −0.00 | 0.00 | 0.00 | 0.00 | −0.00 | −0.00 | −0.00 | 0.00 | −0.00 | 0.00 |
| $f_{U,2015}$ | −0.00 | 0.00 | −0.00 | 0.00 | 0.00 | 0.00 | 0.00 | −0.00 | 0.00 | −0.58 | 0.00 | −0.00 | 0.00 |
| $f_{U,2016}$ | 0.00 | −0.00 | −0.00 | 0.00 | 0.00 | 0.00 | 0.00 | −0.00 | 0.00 | −0.00 | −0.88 | 0.00 | 0.00 |
| $f_{U,2017}$ | −0.00 | 0.00 | −0.00 | 0.00 | 0.00 | 0.00 | −0.00 | −0.00 | −0.00 | 0.00 | −0.00 | −0.44 | 0.00 |
| $f_{U,2018}$ | −0.00 | 0.00 | 0.00 | 0.00 | 0.00 | 0.00 | 0.00 | −0.00 | 0.00 | 0.00 | −0.00 | −0.00 | −0.00 |
| $f_{P,2006}$ | 1.77 | −0.62 | 0.00 | −0.00 | −0.00 | −0.00 | −0.00 | −0.08 | 0.00 | −0.23 | −0.00 | 0.00 | −0.00 |
| $f_{P,2007}$ | 0.00 | 1.80 | −0.62 | 0.00 | −0.00 | −0.00 | −0.00 | −0.10 | −0.00 | 0.00 | −0.00 | −0.00 | −0.00 |
| $f_{P,2008}$ | 0.00 | 0.08 | 1.09 | −0.51 | 0.02 | −0.04 | −0.00 | −0.00 | 0.00 | −0.00 | 0.00 | −0.00 | −0.00 |
| $f_{P,2009}$ | 0.00 | −0.00 | 0.00 | 1.33 | −0.86 | 0.13 | −0.06 | −0.00 | 0.00 | −0.00 | −0.00 | −0.00 | 0.00 |
| $f_{P,2010}$ | 0.00 | 0.00 | 0.00 | −0.00 | 1.69 | −1.15 | 0.23 | −0.10 | −0.00 | 0.00 | −0.00 | 0.00 | −0.00 |
| $f_{P,2011}$ | 0.00 | −0.00 | 0.00 | −0.00 | 0.00 | 2.62 | −1.30 | 0.15 | −0.00 | −0.00 | −0.00 | −0.00 | −0.00 |
| $f_{P,2012}$ | 0.00 | 0.00 | 0.00 | −0.00 | −0.00 | 0.25 | 3.47 | −1.33 | 0.04 | −0.00 | 0.00 | −0.00 | −0.00 |
| $f_{P,2013}$ | 0.00 | 0.00 | −0.00 | 0.00 | −0.00 | 0.00 | 0.00 | 4.15 | −1.17 | 0.05 | −0.16 | 0.00 | −0.00 |
| $f_{P,2014}$ | 0.00 | 0.00 | 0.00 | −0.00 | 0.00 | −0.00 | 0.00 | 0.00 | 3.06 | −1.30 | 0.38 | −0.04 | 0.00 |
| $f_{P,2015}$ | −0.00 | 0.00 | −0.00 | 0.00 | 0.00 | 0.00 | 0.00 | −0.04 | 0.11 | 3.32 | −1.33 | 0.22 | −0.00 |
| $f_{P,2016}$ | 0.00 | 0.00 | 0.00 | 0.00 | 0.00 | 0.00 | 0.00 | −0.00 | 0.00 | 0.03 | 3.30 | −1.25 | 0.44 |
| $f_{P,2017}$ | −0.00 | 0.00 | 0.00 | −0.06 | 0.07 | −0.00 | −0.00 | −0.00 | 0.00 | −0.00 | 0.04 | 3.36 | −1.49 |
| $f_{P,2018}$ | 0.00 | 0.00 | 0.00 | −0.07 | 0.04 | −0.02 | 0.00 | 0.00 | −0.00 | 0.00 | −0.00 | 0.18 | 3.27 |

Table 3: LASSO regression correlation coefficients. We find that $R_0$ for any given year is consistently best predicted by $f_P$ for the same year. For years where the regression allows for more than one nonzero covariate, the $f_P$ from the previous year or $f_U$ of the same year can contribute to the fit. Coefficients for other statistics are not shown since they are equal to zero or negligible.

Then, assuming it is easy to calculate $q'(\vec{\phi}_i)$, we can use all the samples from all iterations in a new formulation for expected values:

$$E[f] = \frac{\sum_{i\in S_1} f(\vec{\phi}_i)c'_i + \sum_{i\in S_2} f(\vec{\phi}_i)c'_i + \cdots + \sum_{i\in S_l} f(\vec{\phi}_i)c'_i}{\sum_{i\in S_1} c'_i + \sum_{i\in S_2} c'_i + \cdots + \sum_{i\in S_l} c'_i} \tag{19}$$

$$c'_i = L(\vec{s} \mid \vec{\phi}_i)\Pi(\vec{\phi}_i)/q'(\vec{\phi}_i) \tag{20}$$

##### 3.4.1 IMIS Algorithm

In our implementation of IMIS, we use summations of uniform distributions as our sampling distributions $q'(\phi)$. The overall structure of the algorithm is shown as a procedure in Algorithm 2. In general, when calibrating to longitudinal data from Thiès, we constrain the dimensions of $\phi$ corresponding to the earlier years of the data first. This allows us to use the causality of transmission dynamics to more efficiently converge on a reasonable parameterization. Details are provided in the following section.

##### 3.4.2 Using IMIS to Reduce Dimensionality of Calibration Procedure

Given that our model must use sequential summary statistics that are temporally correlated, we must evaluate joint likelihoods for all of the statistics over all years. IMIS provides one natural way to deal with these constraints. The algorithm is flexible enough to allow a large number of parameters, and although it requires an exponentially large number of stochastic replicates even when paired with kernel density likelihoods, its iterative aspect allows us to perform dimensional reduction in summary-statistics space to make the KDE more tractable. Taking advantage of the causality of the simulations, we first constrain the parameters for the first few years of data, and then iteratively constrain the latter years. Our method is as follows for a parameterization $\vec{\phi}$ encompassing $M$ summary statistics:

**Algorithm 2** Algorithm for IMIS procedure

**Input:** Provide initial conditions for sampling distribution PDF $q'(\vec{\phi})$. $\vec{\mu}_k$ is a vector for specifying the parameterization of $q_k(\vec{\phi}) = u(\vec{\phi}, \vec{\mu}_k)$, i.e. the $k$th additive component of $q'(\vec{\phi})$ (see Eq. 16). The list $M_\mu = \{\vec{\mu}_k\}$ can then be used to specify $q'(\vec{\phi}) = q_{multi}(\vec{\phi}, \{\vec{\mu}_k\}) = u(\vec{\phi}, \vec{\mu}_1) + u(\vec{\phi}, \vec{\mu}_2) + \cdots + u(\vec{\phi}, \vec{\mu}_l)$.

1: **procedure** IMIS
2: $\quad\vec{\mu} \leftarrow$ initDistribution() ▷ Initialize parameters for sampling distribution
3: ▷ $\vec{\mu}$ will specify the first component of the incremental mixture
4: $\quad\phi_{IM} \leftarrow \varnothing$ ▷ Initialize list of samples
5: $\quad L_{IM} \leftarrow \varnothing$ ▷ Initialize list of sample likelihoods
6: $\quad M_\mu \leftarrow \varnothing$
7: $\quad k \leftarrow 0$ ▷ $k$ tracks the current iteration of the IMIS procedure
8: $\quad$**while** not $f_{converged}(M_\mu)$ **do**
9: $\qquad\vec{\mu}_k \leftarrow \vec{\mu}$
10: $\qquad M_\mu \leftarrow f_{append}(M_\mu, \vec{\mu}_k)$ ▷ $f_{append}$ appends the second provided variable to the first variable
11: $\qquad\{\vec{\phi}_i\}_{q_k} \leftarrow g_{sampler}(\vec{\mu}_k)$ ▷ Draw new samples from an estimate of $P(\vec{\Phi} = \vec{\phi} \mid \vec{s})$ defined by $\vec{\mu}_k$
12: ▷ $\{\vec{\phi}_i\}_{q_k}$ is defined in Eq. 18
13: $\qquad$**for** $\{i \in S_k\}$ **do**
14: $\qquad\quad L_i \leftarrow f_{barcode\_model}(\vec{\phi}_i)$ ▷ Evaluate barcode model as in Section 3.2
15: $\qquad$**end for**
16: $\qquad\phi_{IM} \leftarrow f_{append}(\phi_{IM}, \{\vec{\phi}_i\}_{q_k})$ ▷ Update incremental mixture with new samples and corresponding likelihoods
17: $\qquad L_{IM} \leftarrow f_{append}(L_{IM}, \{L_i\}_{q_k})$
18: $\qquad\vec{\mu} \leftarrow f_{estimate\_posterior}(\phi_{IM}, L_{IM}, M_\mu)$ ▷ Estimate posterior using Eq. 19
19: ▷ Use distribution to update sampling parameters
20: $\qquad k \leftarrow k + 1$
21: $\quad$**end while**
22: **end procedure**

1. For the first round of likelihood evaluation, we run the simulation to generate three years of barcodes over a set of samples $\{\vec{\phi}_i\}_{q_k}$ drawn by the function $g_{sampler}$ (Alg. 2 Line 11) from the PDF $q_k$, which is uniform over a domain specified by $\vec{\mu}$ (Line 2). Each sample is run for multiple stochastic replicates.

2. For the KDE, we use a $M$-dimensional kernel with inflated covariance terms such that $\sigma_j$ are reasonably expected to be larger than the target covariance $\sigma_{kernel,j}$ (Eq. 12) because we do not yet have empirical estimates of $\sigma_{replicates,j}$. With this inflated kernel, we calculate the likelihood (Line 14) and the marginal posterior $P(R_{0,1} \mid \vec{s})$ (evaluated inside the function $f_{estimate\_posterior}$ at Line 18).

3. $f_{estimate\_posterior}$ returns a vector specifying a new sampling distribution based on the marginal posterior for the first year, $P(R_{0,1} \mid \vec{s})$ (Line 18). This results in $q_2$, a new uniform PDF with a reduced domain for $R_{0,1}$. $R_{0,2}$ retains its original domain so that we can explore the full range of dynamics in the second year.

4. For the second round of likelihood evaluation, we run the simulations again, this time sampling with the PDF $q_2$. Combining these samples with those from the first iteration, the effective distribution of all samples is $q' = (q_1 + q_2)/2$ (Eq. 16). We generate four years of barcodes for each replicate of each sample.

5. The updated distribution $q'$ constrains the ranges of summary statistics for the first year. Causality implies that the statistics for the second year (now the first unconstrained year) will be the most informative for selecting models with higher likelihoods.

6. We calculate likelihoods with an updated $M$-dimensional kernel. $\sigma_j$ for statistics associated with the most recently constrained year (currently year 1) are now estimated according to Equation 12 since weighted summary statistics are available to estimate $\sigma_{replicates,j}$.

7. We update the sampling distribution to $q' = q_1 + q_2 + q_3$, specifying $q_3$ based on $P(R_{0,2} \mid \vec{s})$, similarly to step 3.

8. We repeat steps 4 through 7 above for each subsequent year until all years have been fit. $f_{converged}(M_\mu)$ considers the model to be converged when the domain for the last year has been reduced. Given the procedure

above, the final result should be an estimate of the posterior where the all $M$ $\sigma_j$ terms of the likelihood kernel are specified by Eq. 12.

See Subsection 1.2 in Section 1 above for a discussion of results obtained with this IMIS method.

##### 3.5 Code Optimization for Model Calibration

Since model evaluation requires searching over large parameter spaces, it is paramount that each individual simulation runs quickly, and that the overall procedure is parallelizable. Our choice of an IMIS algorithm is intended to address the latter since it is faster than sequential algorithms like gradient descent, but more efficient than evaluating points on a grid.

We optimize run times by running profilers to identify functions in our Python software that tend to use more process time than others. We then refactor or edit the functions while keeping most of their original functionality. For example, we remove Pandas dataframes from the most frequently called functions in order to reduce overhead, and we replace them with Python dictionaries in order to preserve some of the syntax for referencing values by keys. Since the inception of MGMT, we have been able to obtain a 4 to 6-fold reduction in run times using this workflow.

#### 4 Spatial Connectivity

In this section, we describe in more detail the various spatiotemporal analyses conducted with this molecular barcode data and associated metadata including Global Positioning System (GPS) and time information. We begin with aggregated statistics such as the mean distance between pairs of samples that are genetically identical according to the 24-SNP barcode, and follow with methods for characterizing spatiotemporal correlations. The genetic data consists of 2035 molecular barcodes from Thiès from 2006 to 2018. Barcodes are geolocated by catchment, as self-identified by enrolled subjects, and there are varying numbers of clonal and non-clonal samples in each catchment from year to year. The majority of sites are inside the city limits of Thiès, within roughly 8 km of one another (Figure 11).

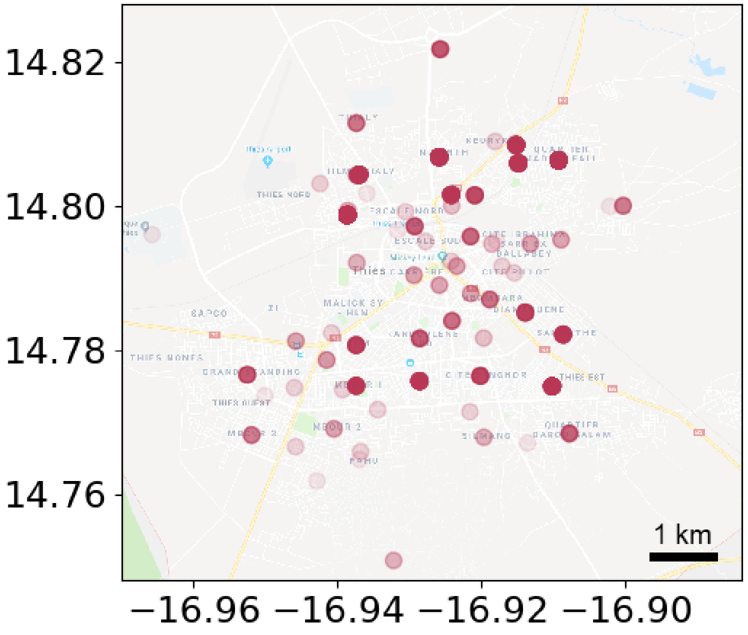


Figure 11: Locations of catchment sites. Proportion of samples from each site denoted by darkness of circle.

#### 4.1 Pairwise Distance Distribution

We find that genetically identical barcodes (clones) also appear geographically close. We find this result to be robust across different ways of representing this relationship.

First, we select for pairs that are at least 14 days and at most 10 km apart. This is to ensure that observations are not from the same infection, and that all distance comparisons remain within the main city borders. We find that clones have a smaller pairwise distance distribution than non-clones, and that the mean and median pairwise distances between bootstrapped subsets of clones are also consistently less than the same for non-clones. The latter holds true when we impose maximum differences of 100 days and 50 days (Figure 12), and also when we allow pairwise comparisons across all years (Figure 13). The largest group of clones is a spatial outlier due to its widespread distribution. We find the above trends to be consistent when we include the group in the analyses, although the effects are less pronounced (Figure 15). On the other hand, the largest group displays an interesting trend, in that it spreads to the majority of neighborhoods in Thiès and is the only group to traverse from the city proper to the satellite catchment over the course of a few years.

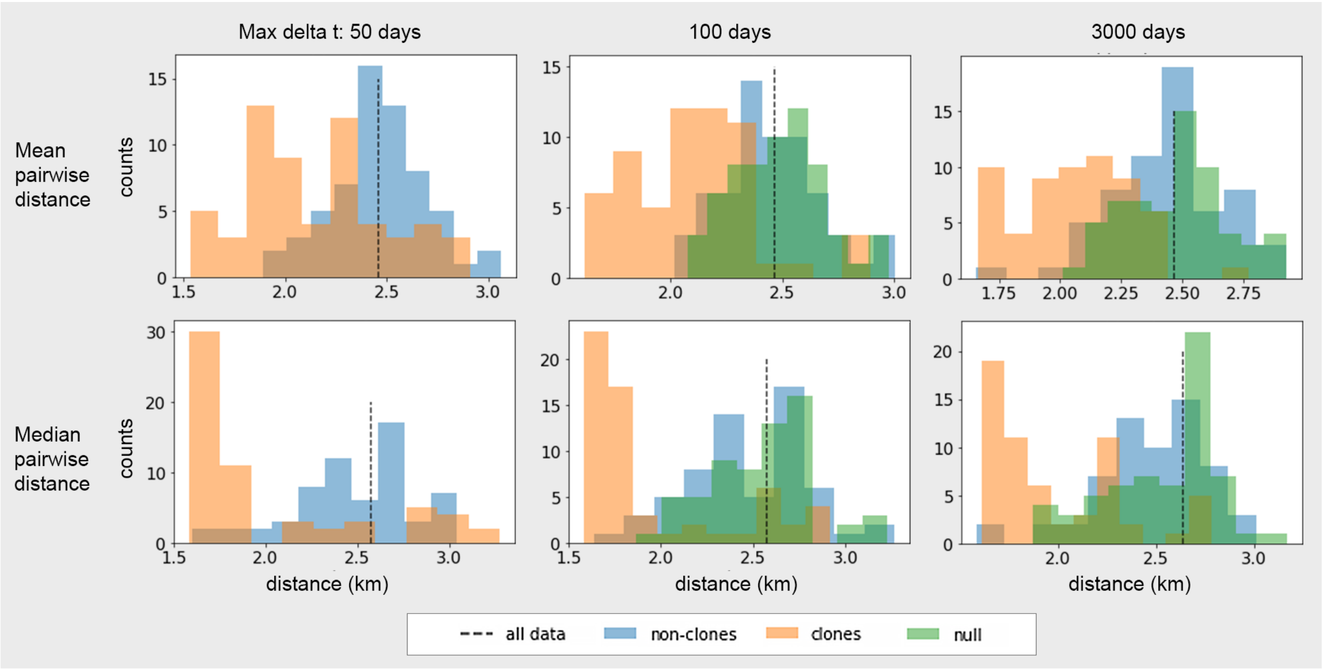


Figure 12: **Distributions of pairwise distances between clones and non-clones.** We estimate the distributions as follows, trying to maintain consistency between different samplings of the data: We group barcodes by strain (sets of barcodes that are identical by state), and then sort the strains by the number of repeated barcodes, from smallest to largest. In order to generate bootstrap samples, we take 35 consecutive barcodes at a time from the grouped and sorted list. This means the first samples include many strains, each with 3 or 4 barcodes each, while the last couple samples are composed almost entirely of the single most repeated strain. For each sample we randomly draw 50 pairs from the 35 barcodes meeting the maximum $\Delta t$ requirements and record the mean and median distances among the pairs. This bootstrap procedure ensures that each strain is equally represented and that each sample reflects the bulk properties of a specific subset of the population that is expected to have shared clustering dynamics (principally due to larger strain groups having a higher likelihood of having propagated out to the maximum distance allowed by the clinic sites). Pairwise distances for non-clones are similarly calculated, except that each grouping of 35 barcodes is randomly selected from the entire set of unique barcodes. The null is constructed by randomly permuting the sample positions while preserving the relative frequency of samples at any given site.

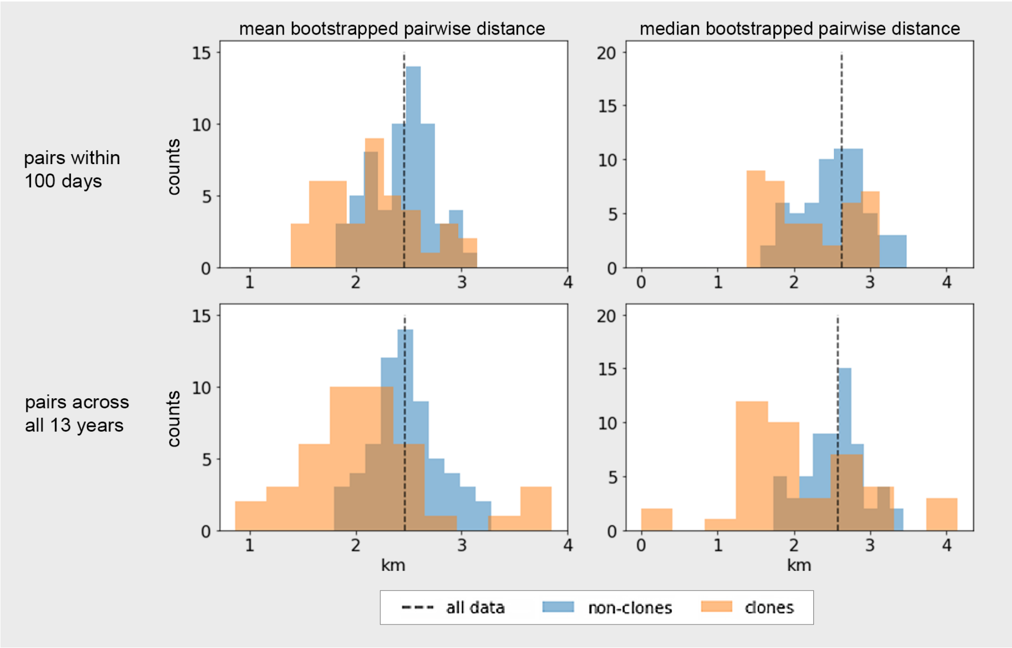


Figure 13: **Distribution of pairwise distances when randomly selecting from all clones.** In this alternate sampling method, we uniformly draw bootstrap samples of 35 barcodes each from the entire set of clonally repeated barcodes (orange histograms), in contrast to the sorted method used in Figure 12. Since this is the same method used for non-clones in Figure 12, the pairwise distributions for non-clones remain unchanged except for stochastic variation and different $\Delta t$ limits (blue histograms). For each sample we draw 50 pairs and record the mean and median pairwise distances. This method results in highly repeated strains having greater influence on the distribution. Nevertheless, we observe a difference between the distributions for clones and non-clones that is in agreement with the alternate method.

Altogether, these sensitivity analyses support the main conclusion that the appearance of persistent clones are spatially correlated. The characteristic distance within the clonal distribution is suggestive about either or both the local chains of transmission within the city or repeated importations to similar neighborhoods from a genetically related pool of parasites.

Similar to foundational work on evaluating power-laws from empirical data [2], we also find the technique of comparing complementary cumulative distributions (cCDF) to be useful for quantitative comparisons and avoids misinterpretation when comparing empirical probability distribution functions. Clones are more closely clustered together in space than non-clones are, and that this relationship reverses for distances greater than $\sim$ 7km when we allow pairwise time-differences greater than 300 days, owing to one particular strain (the strain with the largest number clones) that expands across Thiès over the course of several years, as seen in Figure 14.

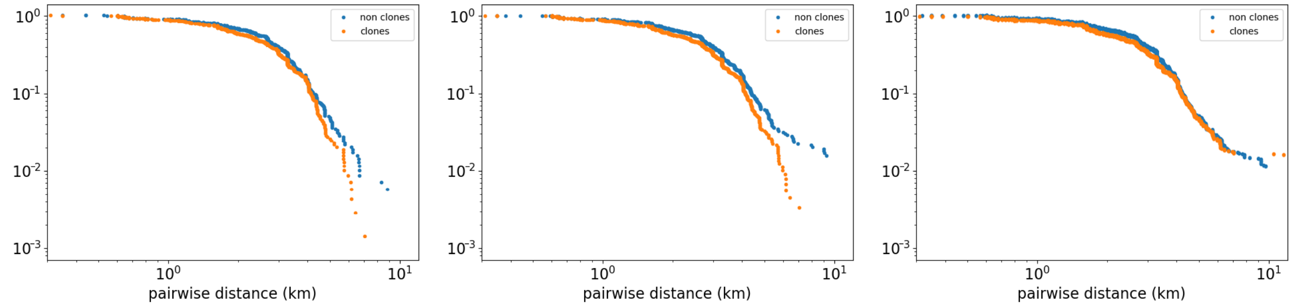


Figure 14: **Complementary cumulative distribution functions (cCDFs) of pairwise distances between clones and between non-clones.** From left to right, we show the distributions corresponding to pairs with maximum time separations of 50 days, 300days, and 3000 days. The cCDF starts with a total probability of 1 at the smallest distances (as indicated by the $y$-axis) and the integrated probability at higher distances is subtracted from the total. Only valid pairs at least 14 days and at most 16 km apart are included. We see that clones are more likely to occur closer together for smaller time separations. For time separations longer than a year, the distribution for clones approaches that for non-clones for distances $>$ 4km, suggesting that as the population mixes over time, spatial correlation among descendants fades for larger distances.

The cCDF of distances between valid pairs of clones (between 14 and 100 days and at most 16 km apart) is markedly different from that of the valid pairs between non-clones. We produce a spatial null model by generating random jumps between the catchment sites assuming that the probability of a jump is proportional to product of the number of data samples at the starting and ending sites. We find that the null cCDF is relatively consistent with the distribution of non-clones. There is a large discrepancy with the cCDF for clones at mid-range distances (1-4 km). This difference is more pronounced and extends to 6 km if we exclude the group of clones with the longest duration, which is a result of the clone group being well-mixed across the city in 2011 (Figure 15).

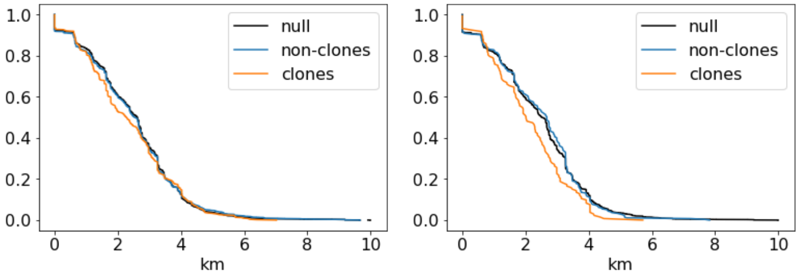


Figure 15: **cCDF for a maximum time separation of 100 days shows significant influence from largest group of clones.** We show the complementary cumulative distribution, indicated by a total probability of 1 on the left. In contrast to Figure 14, the linear axes emphasize differences for mid-range distances. The plot on the left is sampled from all groups of clones, and the plot on the right is sampled from all groups excluding the largest group of clones, which was an outlier based on the number of samples, the extent of its spatial distribution, and the duration of the lineage (five consecutive years). Long-lived clonal expansions have a greater chance of spreading through a population and becoming well-mixed in their latter years, and we expect to see distributions of distances closer to that of a null scenario with no clonal expansion, as demonstrated by the graph on the left.

#### 4.2 Clones also appear to propagate non-randomly

A large percentage of persistent barcodes are correlated in space and time. We leverage principal components to identify these correlations across clones. Whereas correlation coefficients are unable to characterize the slope of the observed trends, the direction and magnitude of principal component vectors concisely capture information about the propagation of clones without requiring a complete covariance matrix. The following is the analysis procedure for computing these relationships:

1. For each set of clones $C_i$ with 3 or more infections, calculate the principal components of the pairwise $\Delta$-distances and $\Delta$-times $(|\Delta\vec{r}|, \Delta t)$ between the clones. We only keep pairs from the same year. Let $\sigma_{i,1}$ and

$\vec{u}_{i,1}$ be the larger singular value and its corresponding principal component unit vector, and let $\sigma_{i,2}$ and $\vec{u}_{i,2}$ be the other singular value and unit vector. Note that $\sigma_{i,1}$ and $\sigma_{i,2}$ should be positive by construction.

2. Create a set of first principal component vectors, $\{\vec{V_i}\} = \{\sigma_{i,1} \cdot \vec{u}_{i,1}\}$ in $(d, \Delta t)$ space, where $d = |\Delta\vec{r}|$.

3. From $\{\vec{V_i}\}$ remove any outlier vectors $\vec{V_i}$ for which either the distance or time elements $V_{i,1}$ or $V_{i,2}$ meet the following criterion: $|V_{i,j}| > r_m V_{m,j}$ where $V_{m,j}$ is the median value of the set of absolute values of vector elements $\{|V_{i,j}|\}$ and $r_m$ is set to 3. For the Thiès data this results in the removal of 3 clonal groups.

4. From the remaining subset of vectors $\{\vec{V_i}\}'$, remove any from clonal groups for which $\sigma_{i,1} < r_\sigma \sigma_{i,2}$, where $r_m$ is set to 10, thus removing approximately half the vectors. This step identifies the clonal groups with strong positive or negative correlations between $|\Delta\vec{r}|$ and $\Delta t$, as well as those that stay in the same location the entire year. For totally random GPS coordinates, these correlations are equally likely to be positive or negative, but for clonal groups that emerge from a specific location due to mechanisms such as the founder effect, these correlations are more likely to be positive.

5. The resulting subset $\{\vec{V_i}\}''$ of vectors is used to analyze the dynamics of persistent clones in space and time. Compute the average vector $\overline{V}$ of the set $\{\vec{V_i}\}''$, and let the correlation slope $s$ be the ratio of its distance and time elements $\overline{V}_1/\overline{V}_2$. (Figure 16)

This method assumes clonal propagation as a simple linear process considering an importation and propagation away from that origin. The local interaction of malaria transmission is significantly more nuanced and detailed; this model does not aim to capture that complexity, but rather suggest the strong correlation of clones appearing in space and time to match our intuition of local chains of transmission and quantitatively demonstrate this behavior using malaria genomic data. The correlation slope $s$ is intended to characterize the spatiotemporal correlation and detect non-random movement of strains. The slope has units of distance over time, but is not strictly a propagation speed. To test our findings, we also formulate several spatial null models for comparison:

1. **Uniform null:** We keep the same phylogenetic structure as the real data, grouping the infections into sets of clones, and letting the number and sizes of the sets match that of the real data. We also keep the same timesteps per infection. We reassign GPS coordinates by randomly placing each infection within the boundaries of Thiès according to a continuous uniform spatial distribution. This null is intended to test the effects of removing spatial correlations between clones.

2. **Discrete uniform null:** We keep the phylogenetic structure and timestamps as above. We reassign GPS locations by randomly placing each infection at the discrete coordinates of a catchment with equal probability.

3. **Discrete weighted null:** This is identical to the discrete uniform null, except that catchments are picked according to probabilities corresponding to their frequency in the real data.

4. **Permuted null:** We keep the phylogenetic structure and the GPS locations of the infections. We scramble up the *order* of infections for each set of clones by randomly permuting the timestamps among each set. This null allows us to test whether the signal we observe in the data is the result of causality.

We find that the correlation slopes estimated from each of these nulls are centered around zero as expected, since any information about spatiotemporal correlations has effectively been erased. We find that the correlation slope estimated from Thiès rejects each of these nulls at better than $p < 10^{-4}$ for the first three scenarios. The final scenario (permuted null) is chosen to have as much spatial structure as possible but still have no time correlation. The corresponding null distribution is influenced by patterns in the data and less constrained than the other nulls, resulting in a rejection at $p < 10^{-3}$ (Figure 18, purple histogram).

We also assess whether we can detect non-random movement of strains using this method. We use two different models of localized movement through Thiès:

1. **Continuous propagation model:** We first simulate propagation as a continuous stochastic process, letting each strain carry out a random walk through the city while keeping the original phylogenetic tree and timestamps. Each displacement vector is calculated with uniform probability in the direction angle and a continuous probability distribution $p(d) \propto d^{-1}$ of selecting any given distance $d$. We calculate a correlation slope for each replicate of the simulation and find that the propagation model assigns greater likelihoods to non-zero slopes (Fig. 18).

2. **Discrete propagation model:** Next, we simulate propagation as a Markov process where parasites are constrained to "jump" from catchment to catchment. The jumping probabilities are specified according to a power law model with an inverse distance dependence: $p = cN_iN_jd^{-1}$, where $d$ is the distance between catchment coordinates and $N_i$ is the number of samples observed at catchment $i$. We also consider an alternate formulation with an exponential jumping probability $p = exp(-d/d_c)$, where $d_c$ is a characteristic jumping distance. As with the continuous model, new positions are assigned while time stamps and phylogenetic relationships remain unmodified. This is a similar spatial null model to previous work using Ebola genetic sequences in Sierra Leone [3]. The discrete model has a similar distribution to the continuous model and also finds non-zero slopes to be more likely, as shown by the light blue histogram in Figure 18.

As described in the main article, we are able to detect non-random movement of strains that are generated using these synethetic models relative to the spatial null models. For parameterizations of these models that are close to a spatial null model, i.e., the exponential kernel in the discrete propagation model being very broad ($d_c >> 700$m), we lose the ability to detect non-random movement (as expected). We conclude that simple methods and metrics such as principal component analysis and a correlation slope can help detect localized transmission and non-random propagation behavior of strains. In future work with more genetic sequences and associated metadata, we believe parsimonious models like these will be useful to detect non-random movement, allow for statistical comparisons to spatial null models, compare to established human movement models, and even parameterize (or fit) movement models with this data for added setting-specific insights.

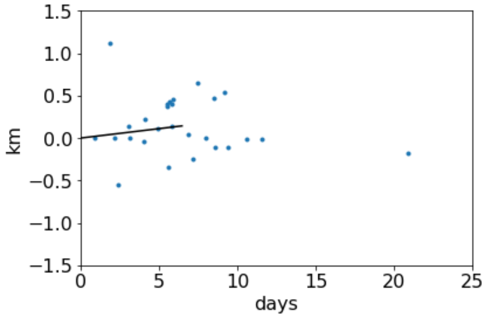


Figure 16: **Calculating the correlation slope from principal component vectors $\{\vec{V}_i\}$.** Each point represents the first principal component vector for a single set of clones from Thiès. The vectors cover a wide range of directions, with some strains showing zero or even negative correlations, but on average the correlations are positive. We compute the average of the vectors (black) and use the slope of the vector to characterize the correlations.

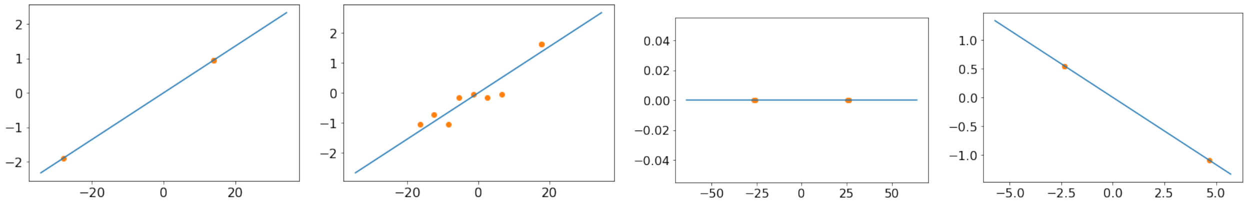

Figure 17: **Principal component analysis (PCA) estimates for representative strains.** The two left plots show the distributions typical of strains that exhibit the type of propagation behavior described in §4.2. Strains that remain in the same site result in a horizontal vector $\vec{u}_{i,1}$. We typically see negative correlations for strains that are first observed in a catchment $C_a$, then a different catchment $C_b$, and finally again in $C_a$.

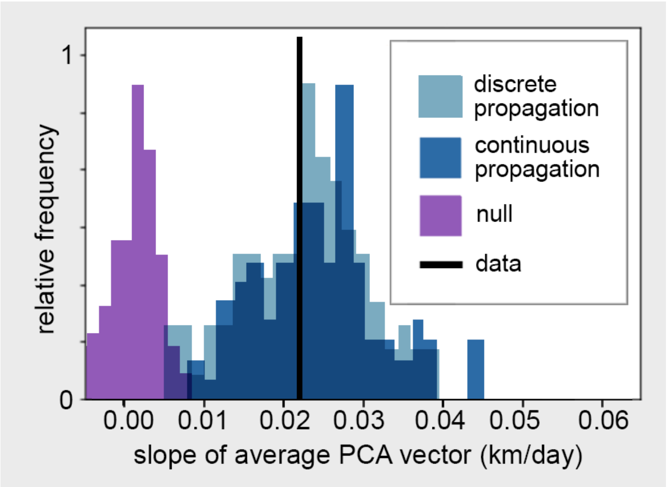


Figure 18: **Distributions of correlation slopes generated from different models.** The solid line indicates the correlation slope estimated from the Thiès data. We compare this to a variety of distributions generated from different models. The permuted null model shown in purple is centered at zero and strongly rejected by the data. The continuous propagation model and discrete propagation model both assign their highest likelihoods to non-zero correlation slopes.

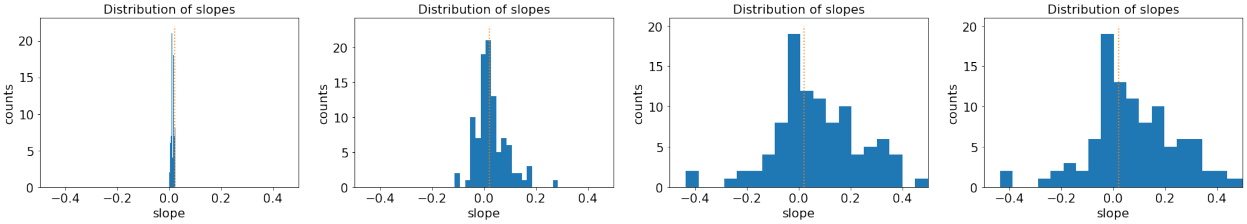


Figure 19: **The exponential discrete propagation model can be varied to generate correlation slopes for different representations of localized transmission in Thiès.** The leftmost plot shows the distribution of slopes derived from simulations with an exponential kernel $d_c$ of 7 meters per day. The other plots show the distributions for $d_c$ = 700 m, 70 km, and 7000 km, respectively from left to right. The last two are intended to roughly model the effects of having a geographically homogeneous population. For a Bayesian interpretation, the most likely parameterization of the propagation model is with $d_c \sim 700$ m. For a frequentist interpretation, the empirical slope strongly rejects the nulls while not specifying a particular propagation model.

### References


[1] RF Daniels, et al., Modeling malaria genomics reveals transmission decline and rebound in senegal. *Proceedings of the National Academy of Sciences* **112**, 7067–7072 (2015).

[2] A Clauset, CR Shalizi, ME Newman, Power-law distributions in empirical data. *SIAM review* **51**, 661–703 (2009).

[3] KB Gustafson, JL Proctor, Identifying spatio-temporal dynamics of ebola in sierra leone using virus genomes. *Journal of the Royal Society* **14** (2017).
